# Physiological variability in key Alzheimer’s biomarkers in amyloid-positive clinical trial cohorts and the mechanistic basis of biomarker ratios

**DOI:** 10.64898/2026.09.18.26363412

**Authors:** Eve Tasiudi, David Hawellek, Eduardo Aponte, Matteo Tonietto, Holly Soares, Cheikh Diack, Antoine Soubret, Tony Kam-Thong, Benjamin Ribba, Marcelo Boareto

## Abstract

**Background:** Protein biomarkers in cerebrospinal fluid (CSF) and plasma have established themselves as essential tools for the diagnosis of neurological disorders and for disease monitoring, thanks to their accuracy and clinical validity. Outside their original intended context of use, protein biomarkers are increasingly used in clinical trials to assess the effects of novel therapies on brain biology. Physiological differences across individuals — such as CSF or plasma volume and elimination kinetics — also contribute to biomarker variability. A deeper understanding of these sources of variability is essential to further improve biomarker interpretation, particularly in clinical trials where populations are by design more homogeneous than in real-world diagnostic settings. This study aimed to quantify the contributions of physiological factors to inter-individual biomarker variability, and to identify the mechanistic basis for why ratio-based normalization can lead to enhanced biomarker performance.

**Methods:** Using a mechanistic kinetic framework and paired CSF and plasma baseline data from four Phase III clinical trials (GRADUATE I and II, CREAD and CREAD2), we quantified physiological and neurobiological contributions (hereafter, physiological and neurobiological variability) to inter-individual variability of commonly used AD biomarkers (Aβ40, Aβ42, p-tau181, t-tau, NfL, GFAP, sTREM2, YKL-40), and evaluated the conditions under which ratio-based normalization can reduce physiological variability.

**Results:** In these amyloid-positive clinical-trial populations, a large fraction of inter-individual variability could be attributed to physiological variability. While normalizing biomarker values by Aβ40 or Aβ42 reduced physiological variability for some biomarkers, the effects of the normalization were compartment-, and cohort-dependent. Using our framework, we identified two conditions under which ratio-based normalization is most likely to improve biomarker performance: (1) the target and reference biomarkers strongly share physiological variability, and (2) they maintain independent neurobiological variability. These conditions provide a mechanistic explanation for why Aβ-based ratios are useful for some biomarkers but are not optimal for others.

**Discussion:** These findings advance our understanding of the sources of biomarker variability in clinical trials and provide a framework for better understanding the mechanistic basis of ratio-based normalization. This work aims to strengthen the utility of using biomarkers by clarifying when and why ratio-based approaches are most informative.

## Introduction

Neurodegenerative disorders are increasing in prevalence worldwide, with Alzheimer’s disease representing the most common cause of dementia^1^. This growing burden has intensified the need for reliable, scalable, and biologically informative tools to support diagnosis, disease staging, patient stratification, and therapeutic research^2–4^. Fluid biomarkers measured in cerebrospinal fluid (CSF) and plasma have become central to this effort because they provide accessible information about neurodegenerative processes in the central nervous system (CNS).

In Alzheimer’s disease, fluid biomarkers have already transformed research and clinical practice by supporting more accurate and biologically grounded diagnosis^5^. Biomarkers related to amyloid pathology, tau pathology, and neuroinflammation - including Aβ42, Aβ40, phosphorylated tau species such as p-tau181 and p-tau217, and glial fibrillary acidic protein (GFAP) - are extensively used to characterize disease biology^1,6^. Their diagnostic importance is increasing further as Alzheimer’s disease diagnosis increasingly depends on early, accurate, and quantitative assessment of Aβ burden and associated pathological processes^6^.

Beyond their diagnostic use, fluid biomarkers also have an important context of use in clinical trials, where they are used to assess target engagement, pharmacodynamic effects, downstream biology, and relationships with clinical outcomes^2–4^. In this context, it is particularly relevant to understand the sources of biomarker variability because clinical trial populations are selected using defined inclusion criteria and are often enriched for a specific diagnosis, pathology, or disease stage^3–7^. As a result, participants may be relatively similar with respect to the neurodegenerative disease process under study. When disease-related heterogeneity is constrained in this way, the relative contribution of non-disease-related physiological differences to inter-individual biomarker variability may become more apparent. For CNS-derived proteins, measured concentrations reflect not only production or release within the CNS, but also movement through CSF and plasma compartments and eventual clearance^8–13^. Differences in biomarker transport, distribution, and clearance between individuals may therefore contribute to measured concentrations^14–16^. As a result, the measured concentration reflects the interplay between CNS production/release, i.e. neurobiological processes, and the physiological processes that govern distribution and absorption/clearance. This raises a central question: to what extent do inter-individual differences in biomarker concentrations reflect differences in CNS production/release versus differences in physiology that modulate biomarker handling?

Several approaches have been used to account for physiological or non-disease-related variability in fluid biomarker concentrations. Regression models commonly adjust for proxy covariates such as age, sex, body mass index, and renal function^17–21^. Moreover, prior work has empirically addressed inter-individual variability (the population-level variation associated with systemic differences) in key AD biomarkers14,16,22-25. For example, Karlsson et al. (2025) used a data-driven approach to show that a substantial portion of CSF biomarker variability can be explained by non-disease factors, such as ventricular volume, and proposed reference biomarkers, including Aβ40, to account for this variability and improve the diagnostic performance of key Alzheimer’s disease biomarkers. More recently, Mravinacová et al. (2025) reported that inter-individual variability in brain-derived proteins was largely independent of disease state and showed that statistical adjustment using either median protein levels or correlated protein pairs could enhance diagnostic performance. Together, these studies underscore the importance of considering inter-individual variability and support the potential value of empirical adjustment strategies. However, most existing approaches do not explicitly quantify how much observed variability arises from CNS production or release versus downstream physiology. As a result, a formal mechanistic framework is still needed to disentangle neuropathological from physiological variability and to clarify under which conditions adjustment is beneficial, when it may be unnecessary, and which biomarkers are most affected.

Mechanistic modeling provides a quantitative framework to address this gap. By explicitly representing how CNS production or release interacts with transport, distribution, and clearance processes, such models can decompose observed inter-individual variability into neurobiological and physiological components. Paired CSF and plasma measurements are particularly informative because they allow variability to be assessed across connected biological compartments rather than within a single fluid alone.

Here, we applied this framework to baseline biomarker data from four Phase III Alzheimer’s disease clinical trials: GRADUATE I, GRADUATE II^26^, CREAD, and CREAD2^27^. These trials provide paired CSF and plasma measurements across biomarkers reflecting Alzheimer’s disease pathology, neurodegeneration, and neuroinflammation, including Aβ40, Aβ42, total tau, p-tau181, NfL, GFAP, sTREM2, and YKL-40. Our objectives were to quantify the extent to which inter-individual differences in biomarker concentrations reflect CNS-related production or release versus physiological processes, and to identify when accounting for such variability may improve biomarker interpretation in clinical trials.

## Results

### Mechanistic modeling partitions inter-individual variability into neurobiological and physiological variability

To quantify sources of inter-individual variability (IIV) in biomarker concentrations, we applied a mechanistic kinetic framework to paired baseline CSF and plasma data from the GRADUATE and CREAD Phase III Alzheimer’s disease trial cohorts. These cohorts provide an informative setting because participants were selected using defined clinical and biomarker criteria, thereby constraining some disease-related heterogeneity while preserving inter-individual differences in physiological factors that may influence biomarker transport, distribution, and clearance.

Specifically, we decompose the total IIV into two distinct components: shared neurobiological variability and the remaining variability attributed to compartment-specific physiological sources (hereon called physiological variability). The shared neurobiological variability defines the population-level differences in the production and release rate of specific biomarkers within the CNS into the CSF. Conversely, physiological variability captures the individual differences in compartment properties, i.e. distribution volumes and absorption/clearance rates that govern biomarker transport, dilution and elimination. Crucially, this classification is strictly based on mechanistic compartmental kinetics rather than a distinction between disease and non-disease biology, as physiological parameters can themselves be altered by disease progression^28,29^.

To make this framework explicit, we model the measured biomarker concentration in each compartment as the product of a shared neurobiological production rate (*R*) and a compartment-specific physiological scaling factor. Specifically, the steady-state concentrations in CSF (*Y*_*C*_) and plasma (*Y*_*P*_) can be expressed as *Y*_*C*_ *= a*_*CSF*_ *R* and *Y*_*P*_ *= a*_plasma_ *R*. In this formulation, *R* represents the rate of biomarker production and release from the central nervous system into the CSF, and the physiological factors (*a*_*CSF*_ and *a*_*plasma*_) represent the compartment-specific physiological factors that dictate how the biomarker is handled systemically. As detailed in the Methods, these factors aggregate distribution volumes (*V*) and clearance rates (*k*) such as *a = (k V)*^−1^, as well as potential peripheral biomarker production (Methods).

Crucially, the ability of this framework to partition inter-individual variability relies on the fundamental hypothesis that the neurobiological production (*R*) and the physiological variability factors (*a*_*CSF*_ and *a*_*plasma*_) are uncorrelated across the evaluated population. Building on this framework, we derived the percentage of physiological variability for CSF and plasma (*F*_*C*_ and *F*_*P*_), defined as the proportion of total variability attributable to physiological factors rather than neurobiological factors (see Methods equations 9–13 for details). We applied this framework to paired baseline CSF and plasma measurements for eight commonly used biomarkers: Aβ40, Aβ42, t-tau, p-tau181, NfL, GFAP, sTREM2, and YKL-40 (Figure 1, Supplementary Figure 1).

**Figure 1:**
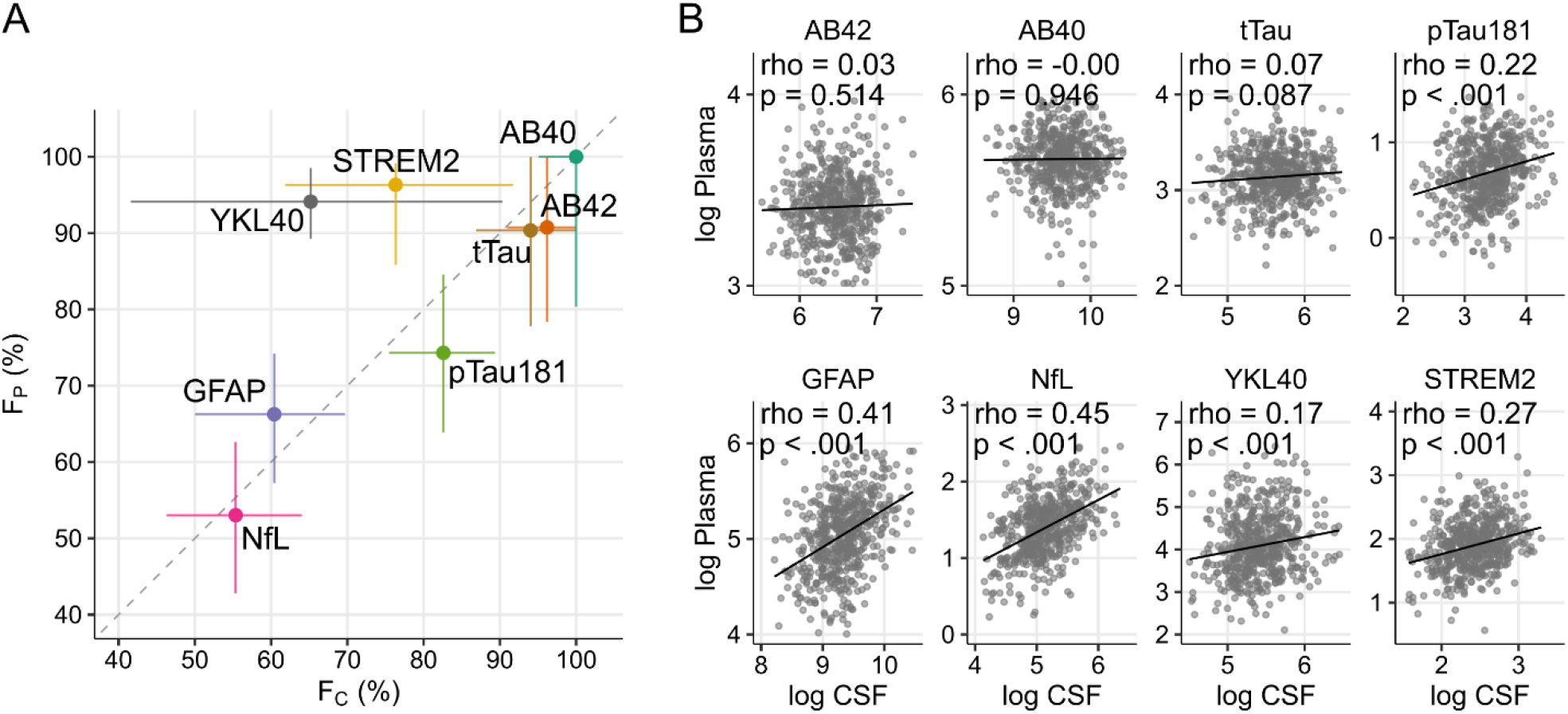
Percentage of physiological variability and CSF-plasma correlations. A) The estimated percentage of inter-individual variability that is attributable to physiological factors in CSF (F_C_, x-axis) and plasma (F_P_, y-axis). B) Scatter plots showing the correlation between paired CSF and plasma concentrations for each biomarker. Each point represents an individual participant. Data: GRADUATE studies. Similar results were obtained for CREAD studies (Supplementary Figure 1). P-values were not corrected for multiple comparisons.

For Aβ40, Aβ42 and t-tau, inter-individual variability was largely attributed to physiological variability in both CSF and plasma. For these biomarkers, *F*_*C*_ and *F*_*P*_ exceeded 90%, indicating that, under the model assumptions and the specific homogeneous clinical trial population, most of IIV differences in concentration were associated with compartment-specific physiology rather than differences in the rate of CNS biomarker release. In contrast, NfL and GFAP exhibited lower fraction of physiological variability in both CSF and plasma (*F*_*C*_, *F*_*P*_ < 70%) (Figure 1A), implying that a larger portion of their IIV reflects differences in the rate of CNS biomarker release. Consistently, we observed higher CSF-plasma correlations (rho>0.36) (Figure 1B, Supplementary Figure 1). Interestingly, for p-tau181, we observed differences between studies, having lower fraction of physiological variability in CREAD (Supplementary Figure 1) as compared to GRADUATE study (Figure 1).

Across most biomarkers, *F*_*C*_ and *F*_*P*_ were of similar magnitude between CSF and plasma, suggesting similar magnitude of physiological variability between CSF and plasma (Figure 1A, Supplementary Table 1). Notable exceptions were sTREM2 and YKL-40, for which the percentage of physiological variability was comparatively lower in CSF than in plasma (Figure 1A). This asymmetry suggests potential peripheral contributions to the plasma levels, adding plasma-specific physiological variability.

### Shared physiological variability contributes to within-compartment correlations

According to our mechanistic framework, correlations among different biomarkers measured within the same compartment, either CSF or plasma, may arise from different sources of shared variability (Methods, equations 16–18). When inter-individual variability is primarily driven by CNS production or release, correlations between biomarkers reflect shared neurobiological processes. In contrast, when a substantial proportion of variability is attributed to compartment-specific physiological variability, biomarkers may also correlate because they are influenced by common processes affecting transport, dilution, or clearance even if their underlying neurobiology is distinct.

Consistent with this framework, biomarkers with the highest estimated contribution of physiological variability, namely Aβ40, Aβ42, and t-tau, showed strong positive correlations within both CSF and plasma (Figure 2). For example, in CSF, t-tau is strongly correlated with Aβ40 (rho=0.65). Notably, these same biomarkers showed minimal CSF-plasma concordance (rho<0.1; Figure 1B).

**Figure 2:**
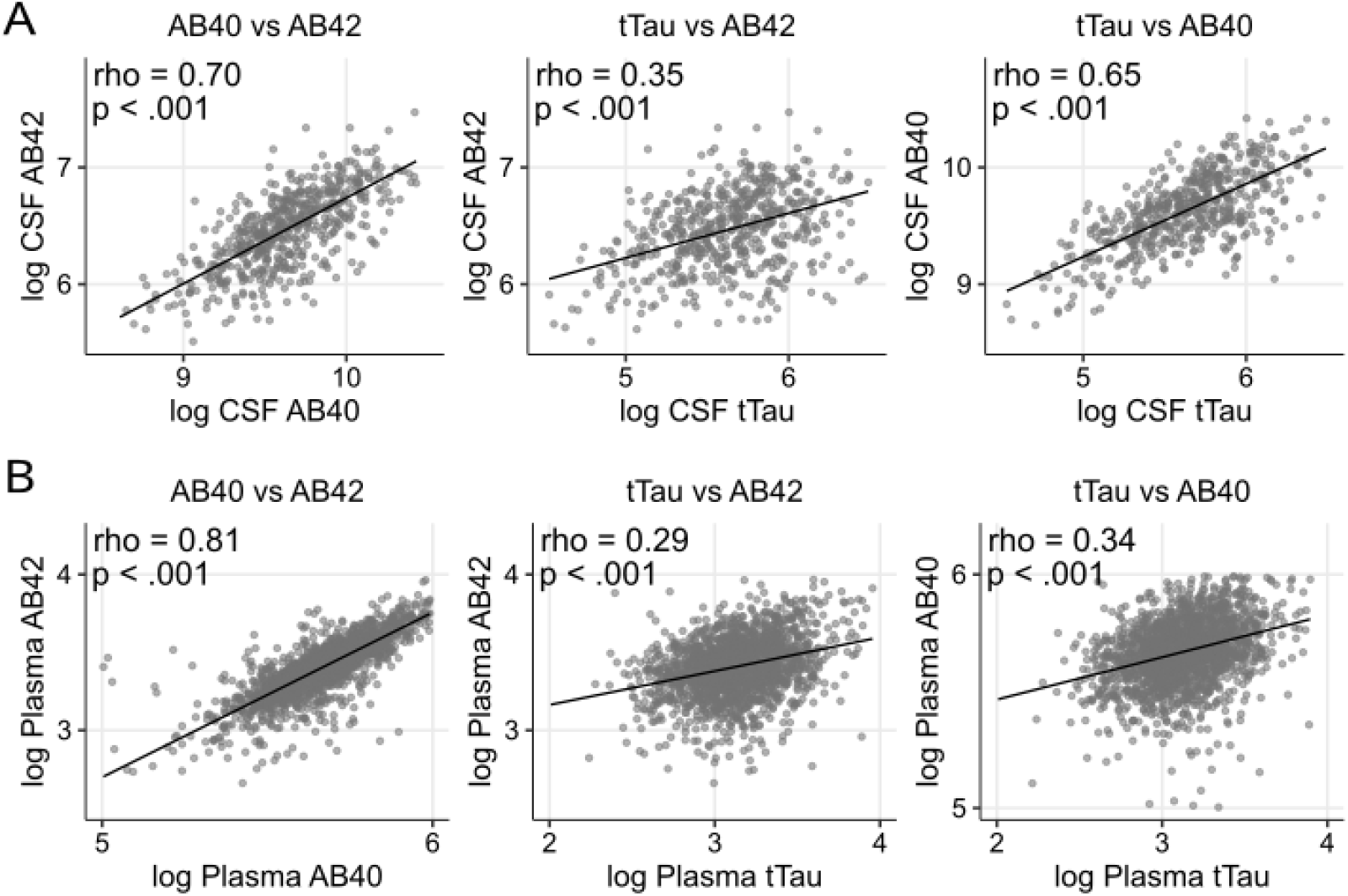
Inter-biomarker correlations in CSF and plasma. Scatter plots showing correlations among Aβ40, Aβ42, and tTau within A) CSF and B) plasma compartments. Each point represents an individual, and the line indicates the linear regression fit, with Spearman rank correlation coefficients (rho) and p-values not corrected for multiple comparisons. Data: GRADUATE studies. Similar results were obtained for CREAD studies (Supplementary Figure 2)

This pattern extends beyond Aβ and t-tau: all analyzed biomarkers displayed positive within-compartment correlations in both CSF and plasma (Figure 3A). To visualize the systemic component of these correlations, we ranked individuals by their average biomarker expression, calculated as the mean Z-score across all biomarkers - where individual concentrations were first standardized relative to the cohort mean and standard deviation - analogous to the approach used by Karlsson et al. (2024). Individuals with higher values for one biomarker tended to have elevated levels across the panel, in both CSF and plasma (Figure 3B). Results were similar in the CREAD cohort (Supplementary Figure 2).

**Figure 3:**
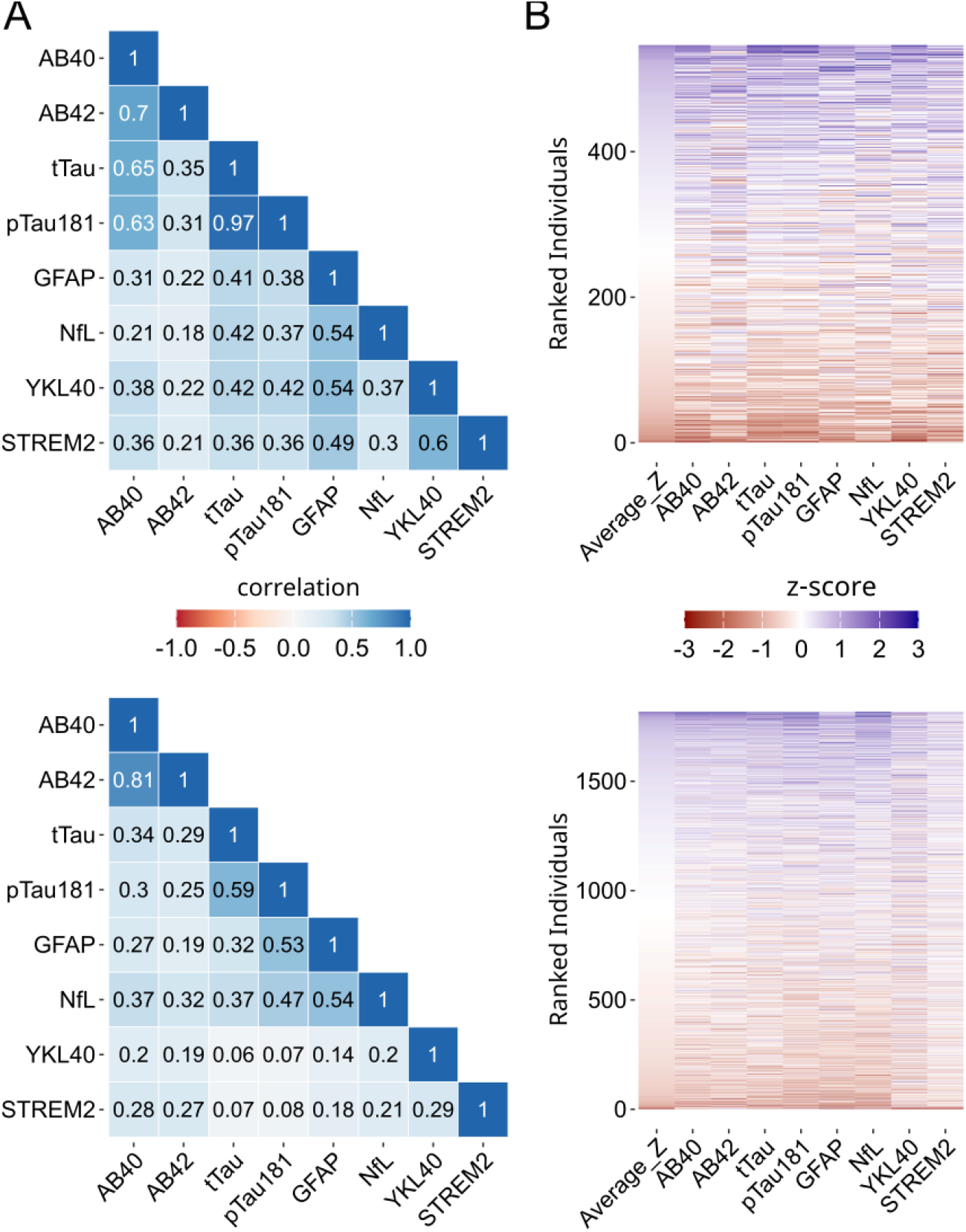
Inter-biomarker correlation and systemic variability in CSF and plasma. A) Heatmaps displaying the Spearman rank correlation coefficients between all pairs of biomarkers measured in CSF and in plasma. The color scale indicates the strength of the correlation, with darker blue representing a stronger positive correlation. B) Heatmaps of Z-scores for each biomarker in CSF and plasma. Individuals (y-axis) are ranked by their average Z-score across all biomarkers. The color scale from red (low Z-score) to purple (high Z-score) shows a consistent pattern where individuals with elevated levels of one biomarker tend to have elevated levels across the entire panel. Data: GRADUATE studies. Similar results were obtained for CREAD studies (Supplementary Figure 3).

### Single biomarkers and biomarker ratios: relationship with physiological parameters and clinical endpoints

We next evaluated whether the model-derived partitioning of inter-individual variability was reflected in associations with measured physiological covariates and clinical endpoints. Specifically, we examined correlations between individual biomarkers, Aβ-normalized biomarker ratios, and two sets of baseline variables: physiological proxy covariates and established Alzheimer’s disease clinical endpoints. Physiological covariates included ventricular volume, used as a proxy for CSF dilution and dynamics; serum creatinine, used as a proxy for renal function and plasma clearance; and body weight, used as a proxy for plasma distribution volume. Clinical endpoints included the Clinical Dementia Rating – Sum of Boxes (CDR-SB), Alzheimer’s Disease Assessment Scale – Cognitive Subscale (ADAS-Cog), and Mini-Mental State Examination (MMSE).

For single biomarkers, those identified by our framework with high physiological variability showed the strongest correlations with physiological covariates. In CSF, Aβ40, Aβ42, and t-tau levels were negatively associated with ventricular volume, whereas in plasma these biomarkers correlated with creatinine (Figure 4A). Conversely, biomarkers with lower estimated percentage of physiological variability - p-tau181, NfL, and GFAP - were more closely related to clinical endpoints (Figure 4A). We then assessed whether empirical normalization to Aβ40 or Aβ42 altered these associations. In CSF, Aβ-normalized ratios for p-tau181 and total tau showed markedly weaker correlations with ventricular volume than the corresponding single biomarkers, together with stronger correlations with CDR-SB, ADAS-Cog, and MMSE (Figure 4). This suggests that, for selected CSF biomarkers, normalization to Aβ species may reduce shared physiological variability and strengthen associations with clinical disease measures. In plasma, the pattern was more heterogeneous. Ratio-based normalization produced only modest reductions in correlations with serum creatinine, and improvements in correlations with clinical endpoints were not consistent between the GRADUATE and CREAD cohorts (Figure 4; Supplementary Figure 4). For GFAP, NfL, sTREM2, and YKL-40, normalization to either Aβ40 or Aβ42 did not provide a clear or consistent benefit in either compartment, with changes in correlations generally small and often discordant between CSF and plasma or between cohorts (Figure 4; Supplementary Figure 4)

**Figure 4:**
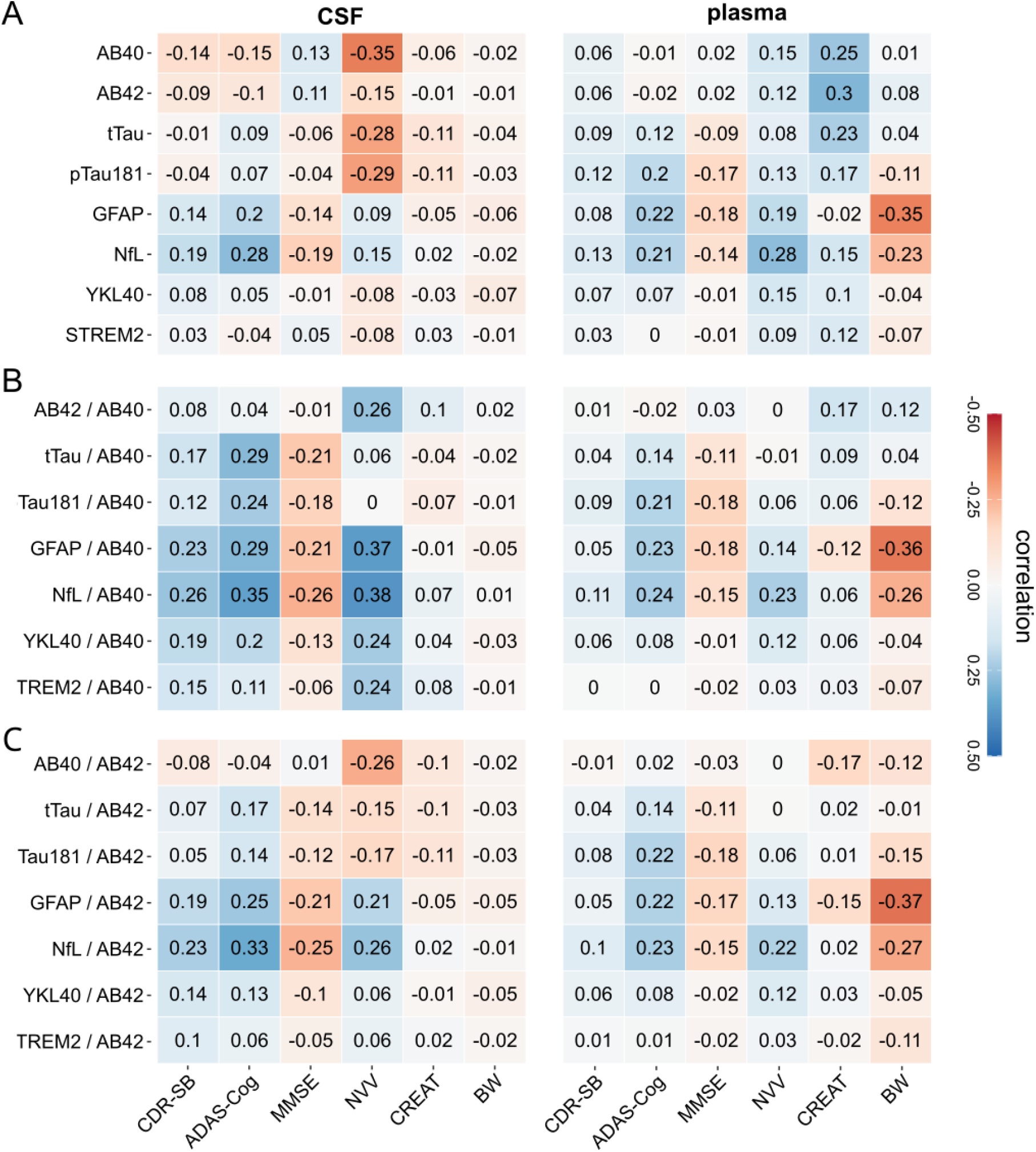
Correlations of single biomarkers and ratios with physiological and clinical variables. Heatmaps showing Spearman rank correlation coefficients between biomarker measurements and clinical endpoints at baseline (CDR-SB, ADAS-Cog, MMSE) and physiological covariates (log-transformed normalized ventricular volume (NVV), creatinine (CREAT), and body weight (BW)). A) CSF and plasma single biomarkers. B) CSF and plasma biomarker ratios normalized to Aβ40. C) CSF and plasma biomarker ratios normalized to Aβ42. The color scale indicates the strength of the correlation, from negative (red) to positive (blue). Data: GRADUATE studies; equivalent results for CREAD studies are shown in Supplementary Figure 4. Number of datapoints per correlation analysis are presented in Supplementary Figure 6.

Together, these findings indicate that the potential value of biomarker normalization depends on the biomarker, compartment, and population under consideration. Aβ-based normalization appeared most informative for selected CSF biomarkers with strong associations to physiological covariates, whereas its benefit was less consistent in plasma and for biomarkers with lower estimated physiological variability. These observations motivated us to ask, from a mechanistic perspective, which properties of the target and reference biomarkers determine whether a ratio should reduce physiological variability and improve biomarker interpretation. We therefore applied our mathematical framework to evaluate how ratio-based normalization redistributes inter-individual variability between CNS production/release and physiological components.

### Mechanistic basis for the utility of biomarker ratios

Biomarker ratios, such as CSF Aβ42/Aβ40 and p-tau181/Aβ42, are widely used because they often improve diagnostic or prognostic performance compared with single biomarkers. These empirical benefits are commonly interpreted as reflecting improved biological specificity, but the mechanisms by which ratio-based normalization changes inter-individual variability are not always explicit. To better understand the patterns observed in Figure 4, we applied our mathematical framework to evaluate how biomarker ratios redistribute inter-individual variability between CNS production/release and physiological variability components.

We first asked under which conditions a ratio reduces the percentage of physiological variability, F_C_ or F_P_, relative to the corresponding single target biomarker. This condition is derived mathematically in Methods Equations 24–30 and defines a region in which such a reduction is theoretically expected, shown as the green region in Figure 5A,B. In CSF, p-tau181 and total tau satisfied this condition when normalized to either Aβ42 or Aβ40. The Aβ42/Aβ40 ratio also satisfied this condition, as did GFAP when normalized to Aβ40 (Figure 5A; Supplementary Figure 5A). In plasma, the predicted benefit was less consistent. In GRADUATE, p-tau181, total tau, and NfL met the criteria for both Aβ denominators, and GFAP when normalized to Aβ40 (Figure 5B). However, these results were not reproduced in the CREAD cohort (Supplementary Figure 5B).

**Figure 5:**
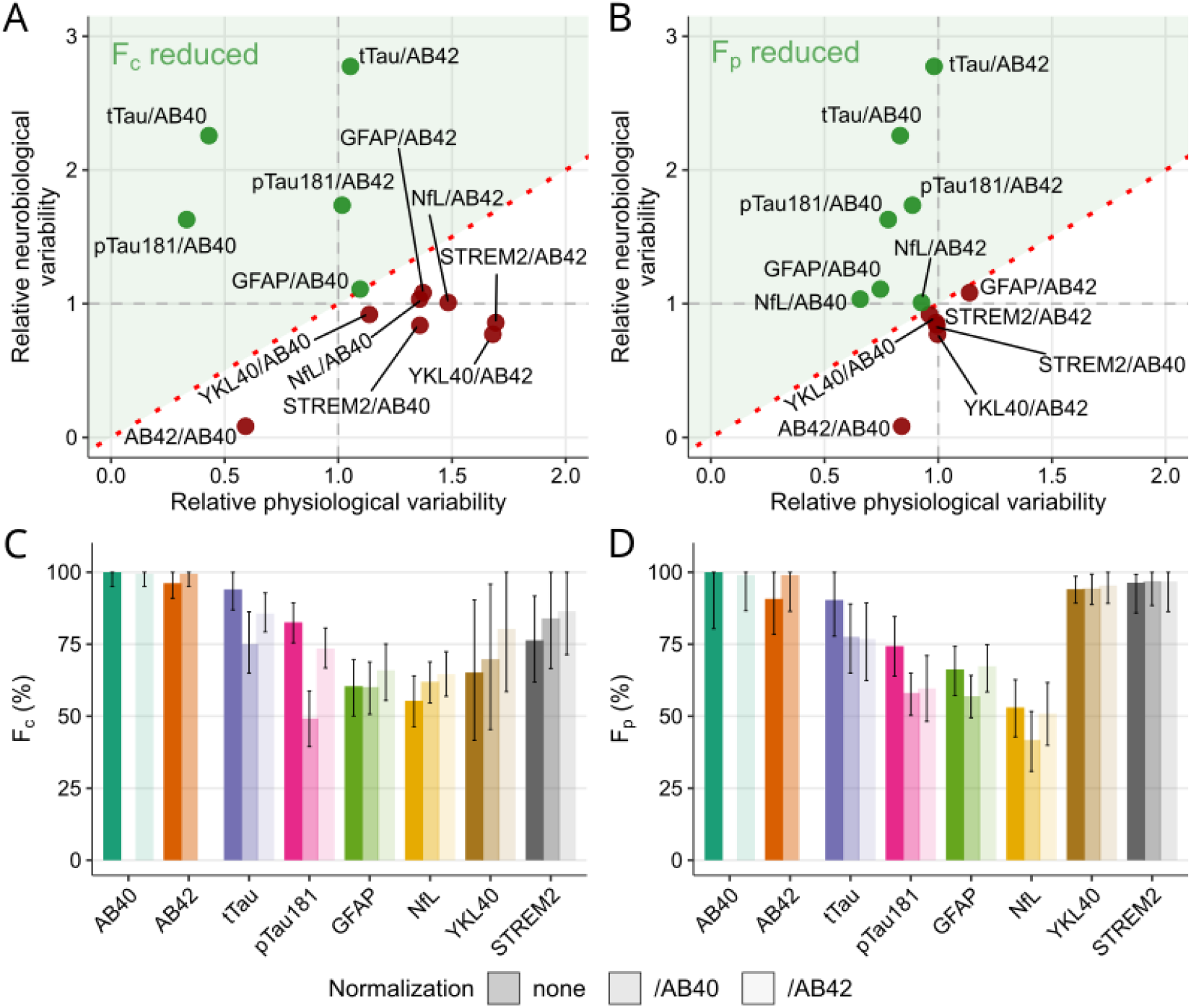
Mechanistic evaluation of biomarker ratios and their impact on physiological variability. (A,B) Scatter plots illustrating the theoretical impact of normalizing target biomarkers by a reference biomarker (Aβ40 or Aβ42) in (A) CSF and (B) plasma. The x-axis represents the estimated relative physiological variability (RPV), i.e., the estimated change in physiological variability of the ratio relative to the single target biomarker; the y-axis represents the estimated relative neurobiological variability (RNV), corresponding change in neurobiological variability of the ratio relative to the single target biomarker. Biomarkers falling within the shaded green region satisfy the mathematical conditions for a reduction in the percentage of physiological variability when the ratio is applied (Methods Equation 30). (C,D) Bar plots comparing the estimated percentage of physiological variability (F_C_ or F_P_) for single biomarker concentrations versus their Aβ40 and Aβ42 ratios in (C) CSF and (D) plasma. Data: GRADUATE studies; equivalent results for CREAD studies are shown in Supplementary Figure 5.

Within this framework, a ratio can reduce the percentage of physiological variability through two non-mutually exclusive mechanisms. First, the ratio can reduce the absolute physiological variability of the target biomarker. This occurs when the target and reference biomarkers share physiological variability— for example, when both are similarly influenced by compartmental volume, dilution, transport, or clearance. In Figure 5A,B, this mechanism is represented on the x-axis, which shows the relative physiological variability of the ratio compared with the single target biomarker. Values below 1 indicate a reduction; for example, a value of 0.4 corresponds to a 60% reduction. Normalization to Aβ40 produced large reductions in relative physiological variability for p-tau181, total tau, and Aβ42 in CSF, with smaller reductions observed in plasma (Figure 5A,B). These model-based predictions are consistent with the empirical correlations shown in Figure 4: in CSF, Aβ40, Aβ42, p-tau181, and total tau showed similar associations with ventricular volume, suggesting a shared physiological component that can be attenuated by ratio-based normalization.

Second, a ratio can reduce the percentage of physiological variability by increasing the relative contribution of CNS production/release variability. This mechanism is represented on the y-axis of Figure 5A,B, where values above 1 indicate that the ratio contains more CNS production/release variability than the single target biomarker. This can occur if the reference biomarker contributes its own neurobiological variability or if the target and reference biomarkers are negatively co-regulated at the CNS production/release level. When the intended purpose of a ratio is to reduce physiological effects, an ideal reference biomarker would share physiology with the target while contributing minimal additional CNS-related variability. Under this idealized scenario, y-axis values would remain close to 1. In CSF, normalization to Aβ40 or Aβ42 increased the relative CNS production/release variability of p-tau181 and total tau, whereas the corresponding values for GFAP, NfL, YKL-40, and sTREM2 were largely unchanged (Figure 5A). A similar pattern was observed in plasma (Figure 5B). The Aβ42/Aβ40 ratio behaved differently, with a reduction in relative CNS production/release variability, consistent with positive co-regulation between Aβ42 and Aβ40 within the framework of the model.

We then quantified the net effect of ratio-based normalization on the percentage of physiological variability, F_C_ and F_P_ (Figure 5C,D; Supplementary Figure 5C,D). Aβ40- or Aβ42-based normalization consistently reduced F_C_ or F_P_ for p-tau181 and total tau, with the largest and most reproducible effect observed for CSF p-tau181 across both GRADUATE and CREAD. In contrast, for GFAP, NfL, sTREM2, and YKL-40, the framework did not predict a robust reduction in physiological variability. This is consistent with the absence of a clear empirical benefit of Aβ-normalized ratios for these biomarkers in Figure 4.

These results provide a mechanistic rationale for when biomarker ratios may be expected to improve biomarker interpretation. The ratio is most likely to reduce physiological variability when the target and reference biomarkers share a substantial physiological component, while remaining sufficiently distinct in their CNS production/release biology. Conversely, ratios may provide limited benefit when the target and reference do not share physiological effects, or when the reference biomarker introduces additional neurobiological variability that changes the biological meaning of the ratio. This distinction is important for interpreting commonly used ratios such as p-tau181/Aβ42, which may reflect both attenuation of shared physiological and integration of biological information from both p-tau181 and Aβ42.

## Discussion

In this study, we developed a mathematical framework to partition inter-individual variability in fluid biomarker concentrations into two model-derived components: variability in CNS production/release and variability in physiology. We applied this framework to paired baseline CSF and plasma data from two independent sets of large Phase III Alzheimer’s disease clinical trials: GRADUATE I/II^26^ and CREAD/CREAD2^27^. These trial cohorts provide an informative setting because participants were selected using defined clinical and biomarker criteria, resulting in populations that are relatively enriched for Alzheimer’s disease pathology and more homogeneous than many observational cohorts. Across these studies, we found that several widely used Alzheimer’s disease biomarkers, including Aβ40, Aβ42, and total tau, had a high model-attributed contribution of physiological variability in both CSF and plasma. In these enriched trial populations, more than 90% of the inter-individual variability for these biomarkers was attributed to compartment-specific physiological variability rather than variability in CNS production/release.

These findings should be interpreted in the context of the populations studied (amyloid positive trial participants) and do not diminish the established diagnostic value of these biomarkers. Rather, they indicate that the relative contribution of physiological variability to measured biomarker concentrations can depend strongly on the setting in which biomarkers are evaluated. In broader populations that include individuals across a wider spectrum of disease stages, the range of CNS production/release may be larger, and the relative contribution of physiological variability may therefore be lower. Conversely, in more homogeneous populations, such as clinical trial cohorts selected for similar pathology or disease stage, variability in CNS production/release may be more constrained. In that context, inter-individual differences in physiological variability — including transport, distribution, dilution, and clearance — may account for a larger share of the remaining cross-sectional variability. Thus, our results are most directly relevant to biomarker interpretation in enriched clinical-trial populations and should not be interpreted as redefining the validated diagnostic use of established biomarkers.

Our findings also have implications for the interpretation of biomarker-biomarker correlations. Within a single compartment, we observed strong positive correlations among several biomarkers. For example, CSF total tau correlated strongly with CSF Aβ40, despite weak CSF-plasma concordance for these biomarkers. Such correlations may reflect shared disease biology in some settings. However, our framework shows that, particularly when biomarkers share physiological variability, within-compartment correlations may also arise because these biomarkers are similarly influenced by compartmental factors such as CSF volume, turnover, transport, or clearance. This provides an additional interpretation for some biomarker associations, especially in enriched clinical-trial populations where disease-related heterogeneity is partly constrained. Therefore, biomarker correlations should be interpreted considering both potential shared neurobiology and potential shared physiological variability.

Adjustments for measured physiological covariates may partially address this issue, but such approaches have limitations^14,16,22–25^. Standard regression models can include proxy covariates such as body weight, serum creatinine, or ventricular volume, and these variables may capture important aspects of plasma distribution, renal function, or CSF dynamics^17–21^. However, these measured covariates are unlikely to fully represent the aggregate physiological processes that influence biomarker concentrations, including fluid turnover, compartmental exchange, tissue distribution, proteolysis, and clearance through multiple pathways. In addition, covariate adjustment can be difficult to interpret if a covariate is itself associated with disease stage or progression. These considerations support the need for complementary mechanistic approaches that explicitly represent how CNS production/release and physiological variability jointly shape measured concentrations.

Our model also provides a mechanistic rationale for the established utility of biomarker ratios. CSF ratios such as Aβ42/Aβ40 and p-tau181/Aβ42 are widely used because they often improve diagnostic or prognostic performance compared with single biomarkers^14,15,30–36^. Our findings support the idea that, in some cases, the denominator of a ratio can act as an empirical reference for shared physiological variability. For example, Aβ40 showed a high model-attributed contribution of physiological variability in both GRADUATE and CREAD. Therefore, using Aβ40 as a reference biomarker may help account for participant-specific physiological effects when the target biomarker and Aβ40 are influenced by similar compartmental processes. This provides a mechanistic explanation for why Aβ-based normalization can improve the interpretability or performance of selected CSF biomarkers.

Importantly, however, ratio-based normalization should not be assumed to be universally beneficial. Within our framework, a ratio is most likely to reduce physiological variability when the target and reference biomarkers share substantial physiological variability while remaining sufficiently distinct in their CNS production/release biology. If the reference biomarker contributes substantial CNS-related variability of its own, the ratio may integrate biological information from both biomarkers rather than simply attenuating shared physiological variability. More broadly, it is important to emphasize that the clinical utility of a biomarker measurement must ultimately be established on a case-by-case basis. The presence of physiological variability, or the current absence of an established normalization strategy, does not preclude a biomarker from yielding important biological insight or meaningful clinical signal. A biomarker that has not yet been optimally normalized still captures disease-relevant variation of considerable value in a clinical trial setting. The framework presented here is therefore intended to refine and contextualize biomarker interpretation, not to serve as a prerequisite for establishing biomarker utility.

These findings are also relevant for the interpretation of pharmacodynamic biomarker effects in clinical trials. Biomarkers are often used to assess target engagement, downstream biological effects, and relationships with clinical outcomes. If measured concentrations are influenced by physiological variability, then treatment-related changes in physiological processes — for example, changes affecting distribution volume, renal function, CSF dynamics, or clearance — could contribute to observed biomarker changes alongside direct effects on CNS disease biology. Analyses of change from baseline partially mitigate this concern, as within-individual baseline measurements inherently anchor observations to each participant’s physiological state, thereby reducing the influence of stable inter-individual physiological differences on longitudinal comparisons. However, this within-individual anchoring does not fully eliminate physiological confounding: if a therapeutic intervention itself alters physiological processes — such as clearance, distribution volume, or CSF dynamics — these treatment-induced physiological changes could drive longitudinal shifts in biomarker concentrations independently of any direct effect on CNS disease biology^25,29,37^. In such cases, an observed change from baseline may reflect a combination of true pharmacodynamic effects and treatment-related physiological perturbations. Appropriately selected biomarker ratios may help reduce shared physiological effects and strengthen interpretation of pharmacodynamic responses. However, this approach requires care. If the reference biomarker is itself affected by the therapeutic intervention, or if the target and reference biomarkers do not share relevant physiological processes, the ratio may be difficult to interpret or may obscure a true biological signal. These considerations highlight the value of using mechanistic frameworks to understand when normalization strategies are likely to improve biomarker interpretation in clinical trials.

Our study has several limitations. First, the framework simplifies the biology of biomarker dynamics. The model assumes that biomarkers are produced or released within the CNS, enter CSF, transfer to plasma, and are ultimately eliminated. Other potential pathways, such as direct brain-to-blood transfer, are not explicitly represented. Second, the framework assumes steady-state conditions, implying a long-term equilibrium between biomarker production/release and clearance. This assumption may be reasonable for slowly progressive neurodegenerative conditions such as Alzheimer’s disease, but may be less appropriate for acute neurological injury or rapidly changing biomarker dynamics. Third, the model assumes that biomarker distributions are sufficiently unimodal within the analyzed population. In populations with strongly bimodal distributions, such as combined healthy and disease groups with clearly separated biomarker levels, the framework may be less suitable or may require extension. Fourth, the model assumes independence between CNS production/release and physiological factors. This assumption may not hold fully in disease settings, because physiological parameters such as brain atrophy, ventricular volume, CSF turnover, or clearance pathways may themselves change with disease progression^17–20,25,28,29^. In addition, this analysis combined two trial programs with different biomarker-based eligibility criteria: GRADUATE required a pathological CSF ptau181/Aβ42 ratio (enriching for tau-positive participants), while CREAD required only abnormal CSF Aβ42^26,27^. These differences in participant selection led to different baseline pathology profiles between cohorts, resulting in discrepancies in physiological variability and ratio performance. Finally, within our framework, the compartment-specific physiological variability term may also capture sources of variability that are not strictly physiological. It is assumed that analytical variability is small relative to total inter-individual variability. This may not hold for all biomarkers, particularly those measured near assay detection limits or at low concentrations. In those cases, model-attributed physiological variability should be interpreted as a term that may include preanalytical and analytical contributions

In conclusion, our study provides a quantitative framework for understanding how CNS production/release and physiological variability jointly contribute to inter-individual variability in CSF and plasma biomarker concentrations. In enriched Alzheimer’s disease clinical-trial populations, where participants are selected for relatively similar disease-defining features, physiological variability can account for a substantial proportion of the inter-individual variability for selected biomarkers. These findings provide a mechanistic perspective that may help refine their interpretation in clinical trials. By clarifying when biomarker levels, biomarker correlations, and biomarker ratios are likely to reflect CNS biology versus shared physiological variability, this framework we hope will support more robust interpretation of fluid biomarker data in Alzheimer’s disease trials and related research settings — while recognizing that the ultimate clinical value of any biomarker, whether normalized or not, remains to be determined through direct evaluation in the relevant disease and context of use.

## Methods

### Mathematical model

To address the challenge of partitioning inter-individual variability (IIV) into neurobiological and physiological variability, we developed a mechanistic framework that estimates the proportion of observed variability attributable to different sources of variability. The framework is grounded in a kinetic model of a brain-derived biomarker spanning production in the central nervous system (CNS), distribution through cerebrospinal fluid (CSF), entry into plasma, and systemic elimination. In this model, the biomarker is produced in the CNS and released into the CSF at rate *R*, with subsequent movement and clearance governed by compartment-specific physiological parameters. It is assumed that biomarker elimination occurs in plasma.

### Model structure and assumptions

The model is based on the following structure and core assumptions:

- **Origin:** The biomarker is produced in the CNS and released into the cerebrospinal fluid (CSF) compartment. It is assumed that the amount of biomarker released per time is constant and referred to as *R*.
- **CSF Dynamics:** The biomarker is assumed to be diluted within an apparent CSF volume (*V*_*c*_) and is cleared from this compartment, primarily via transfer into the blood/plasma, at a rate proportional to its concentration, governed by the rate constant *k*_*p*_. Apparent volume (*V*_*c*_) represents both physical volume and binding to proteins and tissue.
- **Plasma Dynamics:** The biomarker enters the plasma compartment of apparent volume *V*_*p*_ from the CSF. It is then eliminated from plasma through systemic processes (e.g., renal or hepatic clearance) at a rate proportional to its plasma concentration, governed by the rate constant *k*_*e*_. Apparent volume (*V*_*p*_) represents both physical volume and binding to proteins and tissue.
- **Independence between CSF and plasma physiological processes:** Physiological processes contributing to inter-individual variability in plasma and CSF are assumed to be independent.
- **Independence between neurobiological and physiological processes:** Inter-individual variability related to neurobiological processes are assumed to be independent of physiological processes in both plasma and CSF.
- **Steady-State:** For the purpose of analyzing data at baseline, we assume the system is at steady-state, meaning the rates of biomarker production and elimination are balanced.
- **Log-Normal Distribution:** We assume that the model parameters (*R, k*_*p*_, *k*_*e*_, *V*_*c*_, *V*_*p*_) and, consequently, the biomarker concentrations, are log-normally distributed across the population. This is a common and empirically supported assumption for biomarker concentrations, and it allows for a linear representation of the model after log-transformation, simplifying the statistical analysis. Comparison between q-q plots of biomarker concentrations in linear and logarithmic levels support a logarithmic over linear distribution (Suppl. Figures 8–9).
- **Peripheral sources:** Most biomarker production is assumed to occur in the central nervous system, though peripheral sources are also considered. This framework is only appropriate when only a portion of the production originates from peripheral sources; it is not suitable if there is no central nervous system production.

### Derivation of steady-state equations

The change in the amount of biomarker over time for an individual *i* in the CSF (*X*_*ci*_) and plasma (*X*_*pi*_) can be described by the following differential equations:

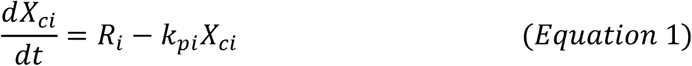

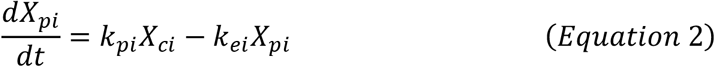

At steady-state, the derivatives are equal to zero (*dX*/*dt* = 0). Since concentration (*Y*) is the amount (*X*) divided by volume (*V*), the steady-state concentrations for an individual *i* are:

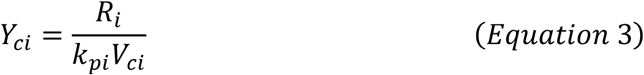

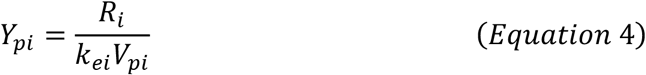

### Decomposition of inter-individual variability (IIV)

To analyze the sources of variability, we log-transform the steady-state equations. The logarithm of the concentration for an individual *i* can be expressed as:

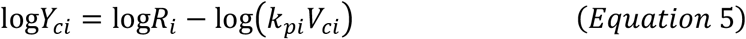

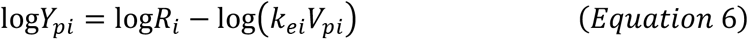

These equations separate neurobiological and physiological terms. The term log*R*_*i*_ is common to both equations. The terms log(*k*_*pi*_*V*_*ci*_) and log(*k*_*ei*_*V*_*pi*_) represent the physiological component specific to the CSF and plasma compartments, respectively.

In the presence of peripheral source of biomarker, Equation 6 can be adapted to take it into account, under the assumption that a fraction of the concentration is from peripheral sources:

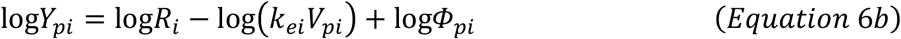

Assuming the rate of CNS biomarker release (*R*) and physiological processes are independent, the total variance of the log-transformed concentrations is the sum of the variances of these components:

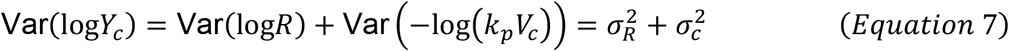

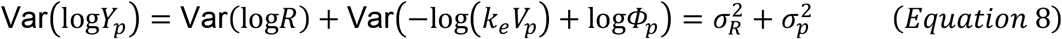

Here, 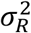 represents the variance related to *R* in the population, while 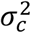 and 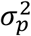 are the variances of the physiological components in CSF and plasma respectively.

## Estimating variance components from paired data

Under the assumption of independence between the rate of CNS biomarker release (*R*) and physiological processes, i.e., Cov(log*R*, log*kV*) = 0, and independence between physiological processes in CSF and plasma, i.e., Cov (−log(*k*_*e*_*V*_*p*_) + log*Φ*_*p*_, −log(*k*_*p*_*V*_*c*_)) = 0, the covariance between the log-transformed CSF and plasma concentrations is driven solely by their shared term (log*R*) which represents the shared neurobiological variability:

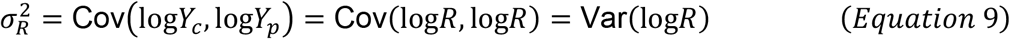

With the total variances and covariance calculated from the data (Equation 7 and 8), we can solve each variance component:

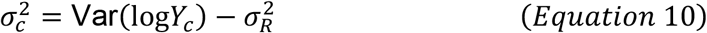

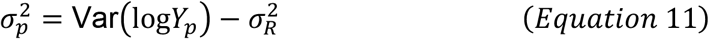

### Estimating the percentage of physiological variability

Finally, we calculate the percentage of total IIV in each compartment that is attributable to physiological variability (*F*_*c*_ and *F*_*p*_):

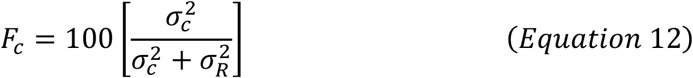

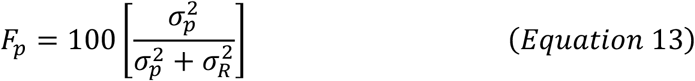

### Estimating the correlation between CSF and plasma

Based on the defined variance components, the theoretical correlation between the log-transformed CSF and plasma concentrations can be directly derived from the model’s structure. The Pearson correlation coefficient, Corr(log*Y*_*c*_, log*Y*_*p*_), is defined as the covariance divided by the product of the standard deviations of the two variables.

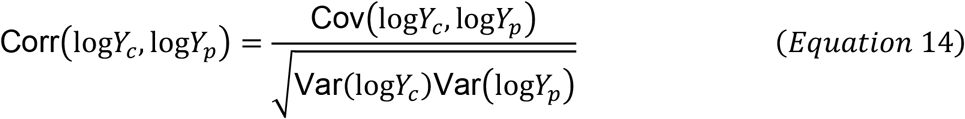

By substituting the terms derived from our mechanistic model (Equations 7–9), we can express the correlation as a function of the neurobiological variance 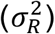 and the compartment-specific physiological variances (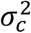and 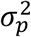):

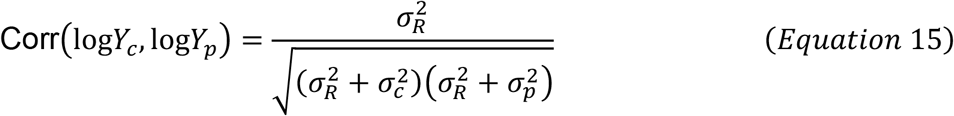

This equation provides a mechanistic link between the sources of inter-individual variability and the observable correlation between CSF and plasma measurements. It shows that a strong correlation is expected only when the variance of the shared central signal 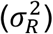 is large relative to the physiological variability in both compartments (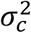 and 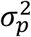). Conversely, if the physiological variability is high in either or both compartments, it will attenuate the correlation.

### Estimating the correlation between two biomarkers in the same compartment

The modeling framework can be further extended to quantify the theoretical correlation between two distinct biomarkers (Biomarker 1 and Biomarker 2) measured within the same biological compartment, such as the CSF. Assuming the independence of CNS production (*R*) and physiological processes, i.e., Cov(log*R*, log*kV*) = 0, the covariance between the log-transformed concentrations of these two biomarkers can be decomposed into two main components:

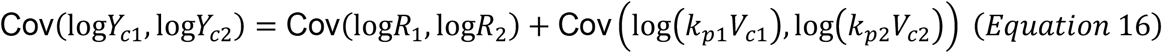

In this equation, the first term, Cov(log*R*_1_, log*R*_2_), reflects the neurobiological relationship between the biomarkers, such as involvement in shared or interacting neuropathological pathways. For simplicity, let’s define: Cov(log*R*_1_, log*R*_2_) = Cov_*R*12_.

The second term, Cov (log(*k*_*p*1_*V*_*c*1_), log(*k*_*p*2_*V*_*c*2_)), represents the covariance of their respective physiological processes. This term captures the extent to which shared physiological processes - like CSF production and turnover rates that influence clearance for both biomarkers - contribute to their correlation. For simplicity, let’s define Cov (log(*k*_*p*1_*V*_*c*1_), log(*k*_*p*2_*V*_*c*2_)) = Cov_*c*12_.

Based on this decomposition, the Pearson correlation coefficient between the two biomarkers in CSF can be expressed as:

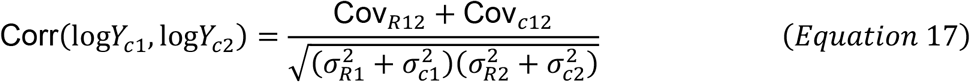

A similar equation can be derived for the correlation between two biomarkers in plasma:

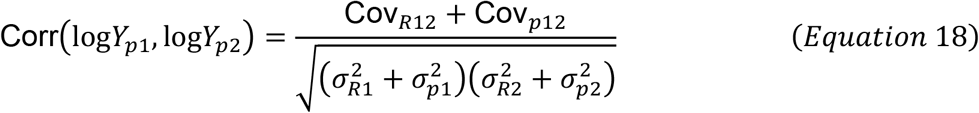

These equations deconstruct the observed correlation into two distinct components. The first part of the numerator quantifies the shared neurobiological variability while the second part quantifies the shared physiological variability. This distinction reveals a critical insight: a correlation between two biomarkers does not, on its own, prove a direct neurobiological link between them. Such a correlation can be driven by shared physiological confounding factors. For instance, if the covariance of the physiological processes (Cov_*c*12_) is much higher than the covariance of their neurobiological processes (Cov_*R*12_), two biomarkers can appear strongly correlated even if their neurobiological pathways are entirely independent.

### Mechanistic interpretation of ratio and necessary conditions

The same framework can be applied to biomarker ratios. Let’s consider a target biomarker (index 1) and a reference marker (index 2), the logarithm transformation of the ratio is defined as:

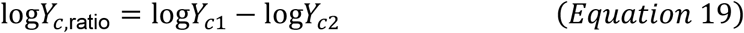

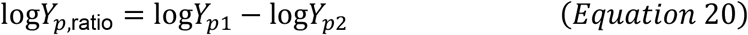

Following the same reasoning of Equations 7–11, the shared neurobiological variance 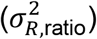, the physiological CSF variance 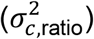, and the physiological plasma variance 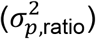 can be estimated as follows:

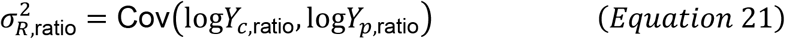

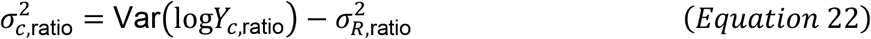

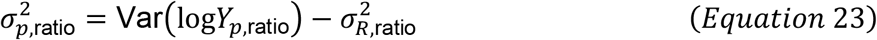

With this, we can define the conditions of when the ratio reduces the percentage of physiological variability as compared to the single biomarker. For the CSF compartment, this condition is defined as:

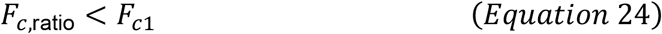

Writing out the physiological fractions, we get:

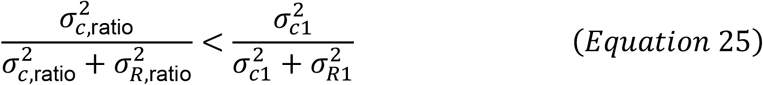

Because variance components are strictly positive, we can cross-multiply and simplify this inequality to:

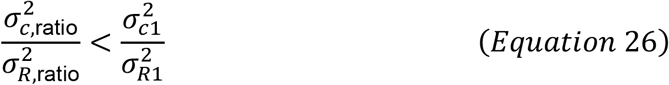

To understand the biological constraints of this inequality, we can expand the ratio variance terms into their individual variances and covariances:

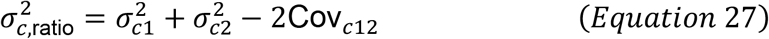

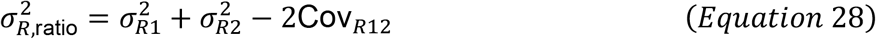

Substituting these back into our inequality reveals the full mechanistic requirement:

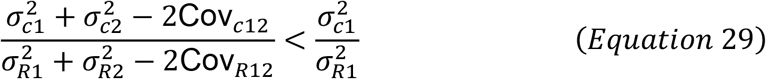

which can be rewritten as:

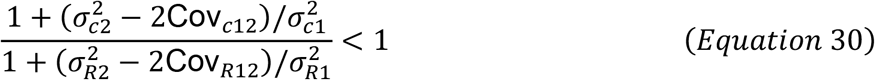

This expanded inequality reveals that for a ratio to successfully lower the fraction of physiological variability while maintaining neurobiological variability, two concurrent biological conditions must be optimized:

- **Maximizing shared physiological variability (minimizing the numerator):** The term 2Cov_*c*12_ must be sufficiently large to offset the additive physiological variability introduced by the reference marker itself 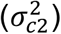. This occurs when both biomarkers are similarly affected by the same physiological processes.
- **Minimizing shared biological variability (maintaining the denominator):** Simultaneously, the term 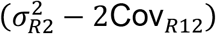 must be minimized. This requires that the reference biomarker contributes negligible neurobiological variability 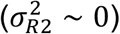 and is not co-regulated with the target biomarker (Cov_*R*12_ ~ 0). Note that the percentage of physiological variability can be reduced by increasing the denominator either when the target and reference biomarker are negatively regulated (Cov_*R*12_ < 0) or the reference biomarker has large neurobiological variability 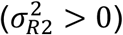. In this case, the neurobiological variability of the ratio is due to the effect of both biomarkers. In contrast, if the target and reference biomarker are positively regulated (Cov_*R*12_ > 0), the ratio can cancel out the neurobiological variability of the target biomarker.

### Study Cohorts

This analysis utilized baseline data from two sets of Phase III multicenter, randomized, double-blind, placebo-controlled clinical trials in early Alzheimer’s disease: the CREAD program (CREAD and CREAD2) and the GRADUATE program (GRADUATE I and GRADUATE II). We leveraged the large participant populations from these trials to enable precise decomposition of physiological and neurobiological contributions.

### GRADUATE Cohorts (Gantenerumab)

The GRADUATE I and GRADUATE II studies evaluated the efficacy and safety of gantenerumab, a subcutaneously administered anti-Aβ monoclonal antibody^26^. These trials enrolled participants aged 50 to 90 years with mild cognitive impairment or mild dementia due to Alzheimer’s disease, with amyloid pathology confirmed by PET or CSF analysis. A total of 2018 participants were randomized to receive either gantenerumab or a placebo.

| Baseline Characteristics - GRADUATE |  |  |
| --- | --- | --- |
| Characteristic | Coverage N (%) | Mean (SD); Median (Min - Max)<sup>†</sup> |
| Age (years) | 2,015 (100%) | 71 (8); 72 (50 - 89) |
| Female | 2,018 (100%) | 1,146 (57%) |
| Education (years) | 2,018 (100%) | 13.3 (4.0); 13.0 (1.0 - 27.0) |
| APOE4 Status | 2,018 (100%) |  |
| 0 copies |  | 672 (33%) |
| 1 copy |  | 1,001 (50%) |
| 2 copies |  | 345 (17%) |
| BMI | 2,001 (99%) | 25.3 (4.4); 24.8 (15.6 - 78.7) |
| CDR-SB | 2,017 (100%) | 3.65 (1.59); 3.50 (0.50 - 11.00) |
| ADAS-Cog | 1,987 (98%) | 28 (7); 28 (6 - 62) |
| MMSE | 2,017 (100%) | 23.6 (3.1); 24.0 (10.0 - 30.0) |
| CSF Aβ42 | 552 (27%) | 674 (227); 649 (186 - 1,757) |
| Plasma Aβ42 | 1,942 (96%) | 30.7 (9.5); 30.1 (3.3 - 254.1) |
| CSF Aβ40 | 550 (27%) | 16,049 (5,261); 15,175 (4,690 - 33,830) |
| Plasma Aβ40 | 1,943 (96%) | 294 (68); 290 (1 - 1,690) |
| CSF pTau181 | 551 (27%) | 32 (14); 29 (9 - 104) |
| Plasma pTau181 | 1,942 (96%) | 2.17 (0.84); 2.03 (0.50 - 11.77) |
| CSF tTau | 551 (27%) | 308 (126); 291 (93 - 819) |
| Plasma tTau | 1,908 (95%) | 24 (7); 23 (1 - 65) |
| CSF NfL | 554 (27%) | 196 (131); 167 (51 - 1,708) |
| Plasma NfL | 1,927 (95%) | 4.37 (2.39); 3.93 (0.46 - 52.30) |
| CSF GFAP | 554 (27%) | 12,613 (8,329); 11,240 (3,700 - 151,300) |
| Plasma GFAP | 1,935 (96%) | 170 (90); 159 (4 - 2,410) |
| CSF YKL-40 | 553 (27%) | 238 (96); 218 (70 - 665) |
| Plasma YKL-40 | 1,853 (92%) | 84 (93); 56 (1 - 1,384) |
| CSF sTREM2 | 550 (27%) | 11.8 (4.1); 11.2 (0.6 - 31.0) |
| Plasma sTREM2 | 1,828 (91%) | 7.22 (3.43); 6.62 (0.00 - 76.45) |

<sup>†</sup> Mean (SD); Median (Min - Max); n (%)

### CREAD Cohorts (Crenezumab)

The CREAD and CREAD2 studies investigated the efficacy and safety of crenezumab, an anti-Aβ monoclonal antibody^27^. The trials enrolled participants between 50 and 85 years of age with a diagnosis of early Alzheimer’s disease (prodromal-to-mild) confirmed by either cerebrospinal fluid (CSF) Aβ42 levels or amyloid positron emission tomography (PET). Key inclusion criteria were a Mini-Mental State Examination (MMSE) score of 22 or higher and a Clinical Dementia Rating Scale-Global Score (CDR-GS) of 0.5 or 1.0. In total, 1611 participants were randomized to receive either 60 mg/kg of crenezumab or a placebo intravenously every four weeks.

| Baseline Characteristics - CREAD |  |  |
| --- | --- | --- |
| Characteristic | Coverage N (%) | Mean (SD); Median (Min - Max)<sup>†</sup> |
| Age (years) | 1,611 (100%) | 71 (8); 72 (50 - 85) |
| Female | 1,611 (100%) | 934 (58%) |
| Education (years) | 1,611 (100%) | 13.3 (3.6); 14.0 (6.0 - 18.0) |
| APOE4 Status | 1,611 (100%) |  |
| 0 copies |  | 495 (31%) |
| 1 copy |  | 826 (51%) |
| 2 copies |  | 290 (18%) |
| BMI | 1,598 (99%) | 25.3 (4.1); 24.9 (13.3 - 46.9) |
| CDR-SB | 1,611 (100%) | 3.78 (1.60); 3.50 (0.50 - 11.00) |
| ADAS-Cog | 1,609 (100%) | 29 (7); 29 (6 - 63) |
| MMSE | 1,611 (100%) | 23.54 (2.91); 24.00 (9.00 - 30.00) |
| CSF Aβ42 | 628 (39%) | 594 (176); 574 (200 - 1,182) |
| Plasma Aβ42 | 798 (50%) | 31.3 (8.3); 30.7 (2.5 - 115.0) |
| CSF Aβ40 | 625 (39%) | 16,563 (5,301); 16,110 (3,530 - 34,810) |
| Plasma Aβ40 | 798 (50%) | 316 (477); 287 (11 - 9,400) |
| CSF pTau181 | 628 (39%) | 34 (17); 32 (8 - 120) |
| Plasma pTau181 | 804 (50%) | 2.30 (0.88); 2.17 (0.52 - 6.29) |
| CSF tTau | 628 (39%) | 345 (155); 319 (80 - 1,220) |
| Plasma tTau | 0 (0%) | NA (NA); NA (Inf - -Inf) |
| CSF NfL | 629 (39%) | 207 (134); 180 (14 - 1,997) |
| Plasma NfL | 784 (49%) | 4.18 (1.81); 3.84 (0.97 - 14.77) |
| CSF GFAP | 641 (40%) | 12,357 (7,736); 11,430 (166 - 124,300) |
| Plasma GFAP | 791 (49%) | 207 (92); 192 (32 - 1,230) |
| CSF YKL-40 | 633 (39%) | 198 (80); 187 (23 - 973) |
| Plasma YKL-40 | 0 (0%) | NA (NA); NA (Inf - -Inf) |
| CSF sTREM2 | 630 (39%) | 10.7 (3.5); 10.4 (0.3 - 28.6) |
| Plasma sTREM2 | 0 (0%) | NA (NA); NA (Inf - -Inf) |

<sup>†</sup> Mean (SD); Median (Min - Max); n (%)

### Data Transformation and Normalization

Baseline biomarker concentrations (CSF and plasma) were log-transformed prior to analysis. Raw linear values for these variables typically exhibit highly skewed distributions; log-transformation normalizes the data. The appropriateness of this log-normal assumption was confirmed using quantile-quantile (Q-Q) plots (Supplementary Figures 8–9). Ventricular volume was normalized according to established methods (Smith et al, 2002).

Cross-sectional associations between biomarker levels, clinical endpoints (CDR-SB, ADAS-Cog, MMSE), and physiological covariates (NVV, CREAT, BW) were evaluated uniformly using Spearman rank correlation (rho). We decided to use a rank-based approach to handle the discrete, bounded nature of the cognitive scoring scales. For statistical consistency across the manuscript, the Spearman metric was maintained for continuous physiological covariates as well; for these log-linear relationships, rank-based and linear coefficients (ie Pearson) yield highly convergent values. The squared coefficient (rho^2) provides the proportion of rank-transformed variance explained.

### Standard Protocol Approvals, Registrations, and Patient Consents

For all cohorts, participants provided written informed consent, and the studies were conducted in accordance with Good Clinical Practice guidelines and the principles of the Declaration of Helsinki. Detailed descriptions of the study protocols have been previously published for the GRADUATE studies (ClinicalTrials.gov Identifier:, NCT03444870, NCT03443973, reference 26) and CREAD studies (ClinicalTrials.gov Identifier: NCT02670083, NCT03114657, reference 27).

### Software

Figure generation and statistical analysis were performed in R version 4.4.1. An interactive web application was developed in R (version 4.4.1) using the shiny framework to calculate and visualize physiological variability in Plasma and CSF. The tool utilizes tidyverse packages for dynamic cohort filtering and automatic logarithmic transformations. Data distributions are evaluated via automated Shapiro-Wilk tests and visual diagnostics (histograms, Q-Q plots) using ggplot2. To ensure statistical reliability, 95% confidence intervals are derived via 1,000 bootstrap iterations.

## Data Availability

For clinical trial studies, qualified researchers may request access to individual patient-level clinical data through a data request platform. At the time of writing, this request platform is Vivli (vivli.org/ourmember/roche/). Up-to-date details on Roche’s Global Policy on the Sharing of Clinical Information and how to request access to related clinical study documents can be found at go.roche.com/data_sharing. Anonymized records for individual patients across more than one data source external to Roche cannot, and should not, be linked because of a potential increase in risk of patient re-identification.

## Acknowledgments

The authors thank all GRADUATE and CREAD trial participants, their families, the site staff, and the trial team, for their time and commitment to the trial, and the following colleagues for their critical feedback on this manuscript: Geoff Kerchner and Nicolas Frey. The authors also thank the Roche Alzheimer’s Disease Data Mart Analytics Team for the creation of the LeAD datamart. This study was sponsored by F. Hoffmann-La Roche Ltd, Basel, Switzerland.

## Supplementary Information

**Supplementary Table 1:**
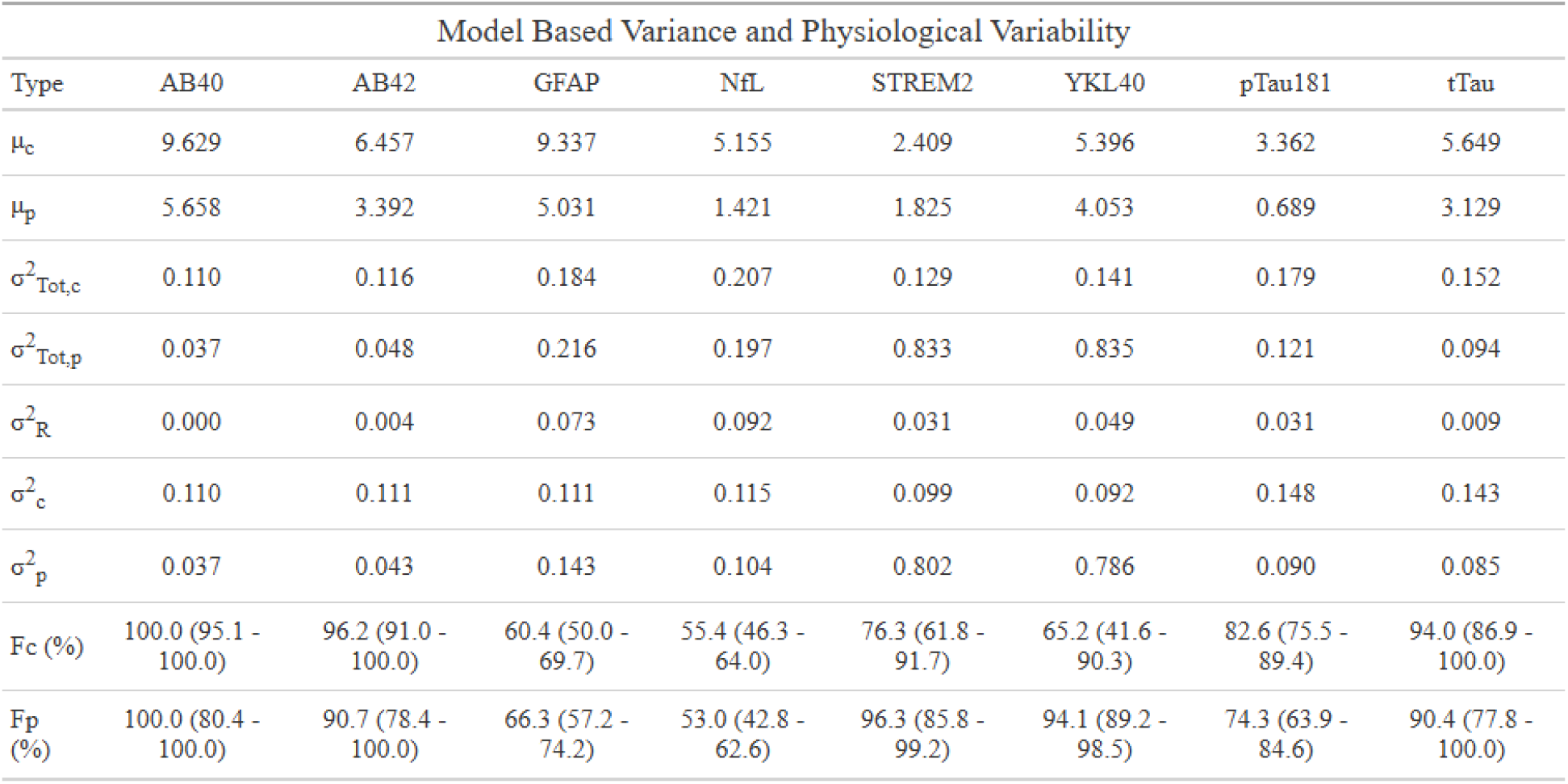
Model based inter-individual variability estimation in CSF and plasma for GRADUATE studies (for CREAD see Supplementary Table 2).

| Model Based Variance and Physiological Variability |  |  |  |  |  |  |  |  |
| --- | --- | --- | --- | --- | --- | --- | --- | --- |
| Type | AB40 | AB42 | GFAP | NfL | STREM2 | YKL40 | pTau181 | tTau |
| $\mu_c$ | 9.629 | 6.457 | 9.337 | 5.155 | 2.409 | 5.396 | 3.362 | 5.649 |
| $\mu_p$ | 5.658 | 3.392 | 5.031 | 1.421 | 1.825 | 4.053 | 0.689 | 3.129 |
| $\sigma^2_{Tot,c}$ | 0.110 | 0.116 | 0.184 | 0.207 | 0.129 | 0.141 | 0.179 | 0.152 |
| $\sigma^2_{Tot,p}$ | 0.037 | 0.048 | 0.216 | 0.197 | 0.833 | 0.835 | 0.121 | 0.094 |
| $\sigma^2_R$ | 0.000 | 0.004 | 0.073 | 0.092 | 0.031 | 0.049 | 0.031 | 0.009 |
| $\sigma^2_c$ | 0.110 | 0.111 | 0.111 | 0.115 | 0.099 | 0.092 | 0.148 | 0.143 |
| $\sigma^2_p$ | 0.037 | 0.043 | 0.143 | 0.104 | 0.802 | 0.786 | 0.090 | 0.085 |
| F<sub>c</sub> (%) | 100.0 (95.1 - 100.0) | 96.2 (91.0 - 100.0) | 60.4 (50.0 - 69.7) | 55.4 (46.3 - 64.0) | 76.3 (61.8 - 91.7) | 65.2 (41.6 - 90.3) | 82.6 (75.5 - 89.4) | 94.0 (86.9 - 100.0) |
| F<sub>p</sub> (%) | 100.0 (80.4 - 100.0) | 90.7 (78.4 - 100.0) | 66.3 (57.2 - 74.2) | 53.0 (42.8 - 62.6) | 96.3 (85.8 - 99.2) | 94.1 (89.2 - 98.5) | 74.3 (63.9 - 84.6) | 90.4 (77.8 - 100.0) |

**Supplementary Table 2:** Same as Supplementary Table 1 for CREAD studies. NA indicates biomarker measurement was not available either in plasma or CSF

| Model Based Variance and Physiological Variability |  |  |  |  |  |  |  |  |
| --- | --- | --- | --- | --- | --- | --- | --- | --- |
| Type | AB40 | AB42 | GFAP | NfL | STREM2 | YKL40 | pTau181 | tTau |
| $\mu_c$ | 9.645 | 6.312 | 9.283 | 5.198 | NA | NA | 3.388 | NA |
| $\mu_p$ | 5.627 | 3.383 | 5.215 | 1.355 | NA | NA | 0.700 | NA |
| $\sigma^2_{Tot,c}$ | 0.117 | 0.096 | 0.250 | 0.242 | NA | NA | 0.268 | NA |
| $\sigma^2_{Tot,p}$ | 0.065 | 0.065 | 0.182 | 0.189 | NA | NA | 0.170 | NA |
| $\sigma^2_R$ | 0.002 | 0.007 | 0.082 | 0.115 | NA | NA | 0.108 | NA |
| $\sigma^2_c$ | 0.115 | 0.090 | 0.168 | 0.127 | NA | NA | 0.160 | NA |
| $\sigma^2_p$ | 0.062 | 0.058 | 0.100 | 0.073 | NA | NA | 0.063 | NA |
| F<sub>c</sub> (%) | 97.9 (91.1 - 100.0) | 93.1 (85.8 - 100.0) | 67.3 (49.8 - 78.7) | 52.3 (41.8 - 61.6) | NA | NA | 59.9 (52.4 - 67.6) | NA |
| F<sub>p</sub> (%) | 96.2 (67.8 - 100.0) | 89.7 (75.4 - 100.0) | 55.1 (39.1 - 69.7) | 38.9 (26.9 - 50.8) | NA | NA | 36.8 (24.6 - 49.7) | NA |

**Supplementary Figure 1.**
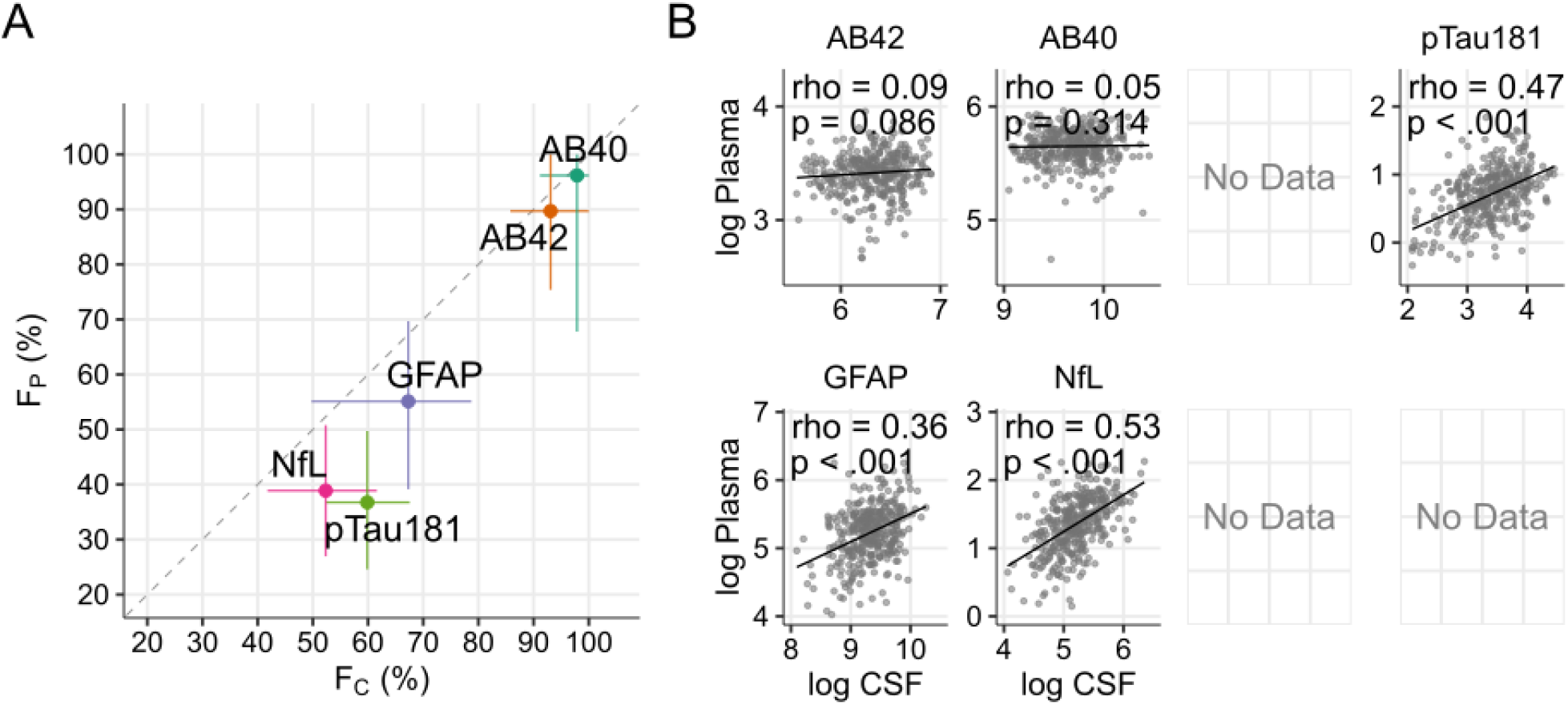
same as Figure 1 for CREAD dataset.

**Supplementary Figure 2:**
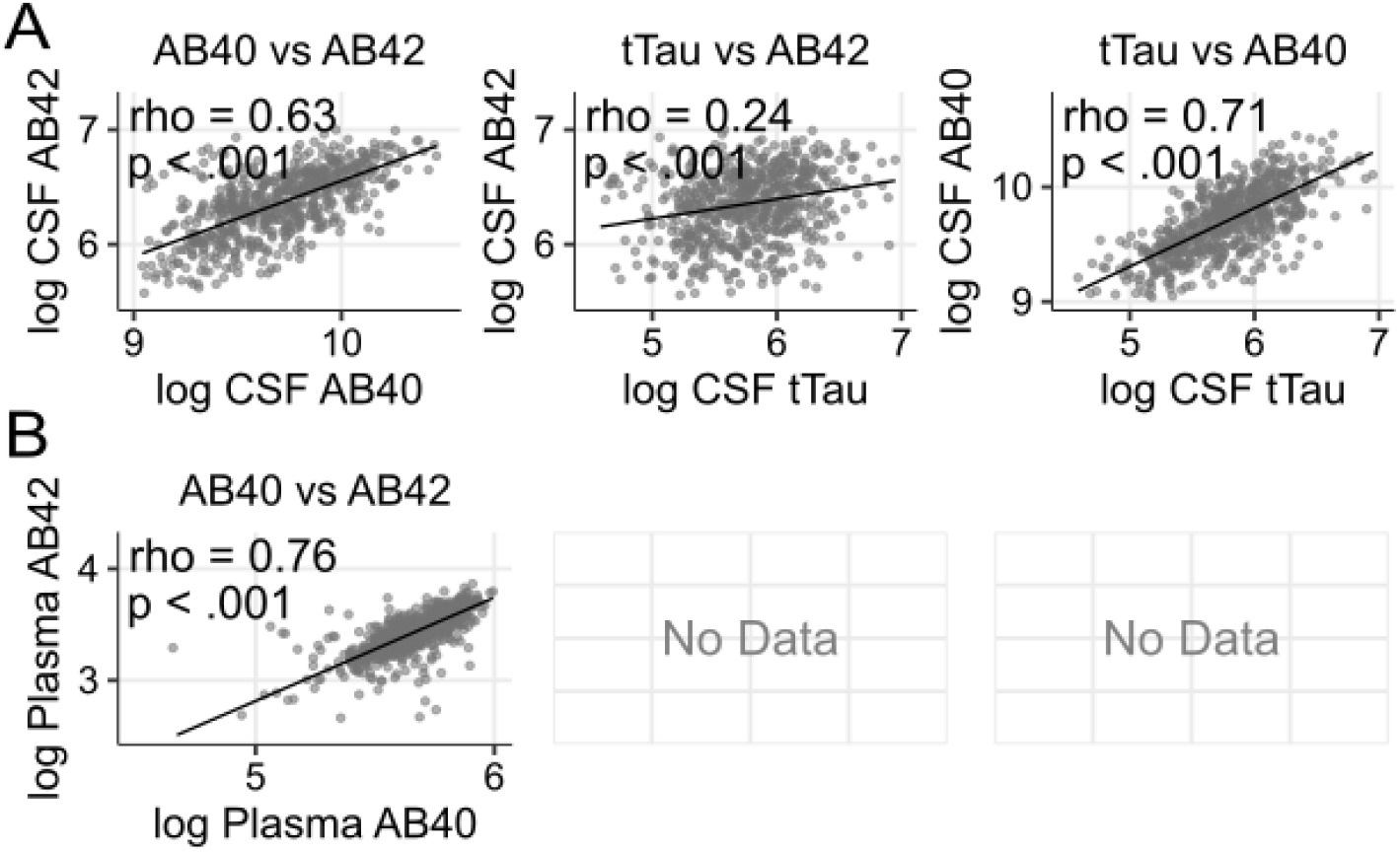
same as Figure 2 for CREAD dataset.

**Supplementary Figure 3:**
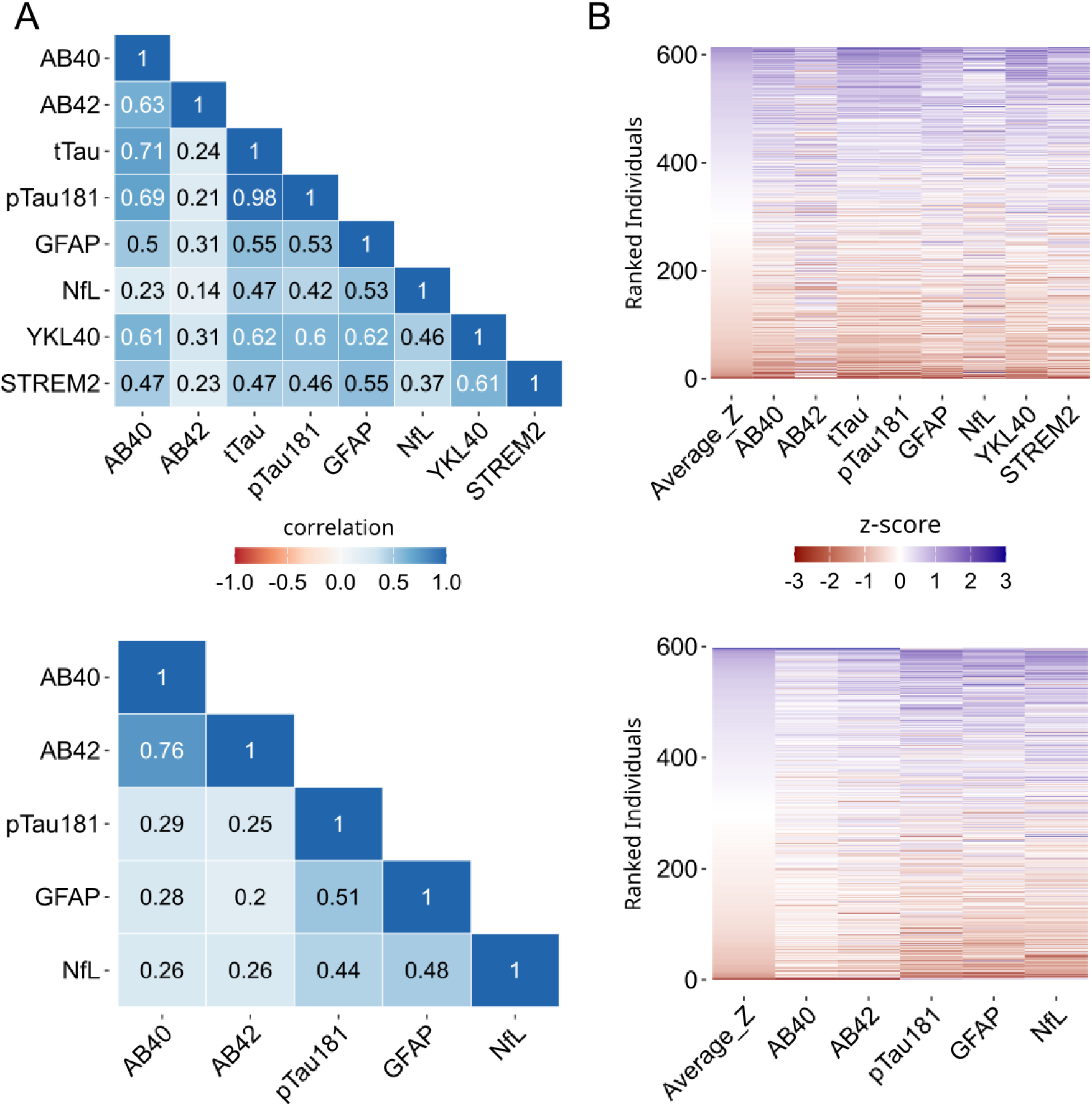
same as Figure 3 for CREAD dataset.

**Supplementary Figure 4:**
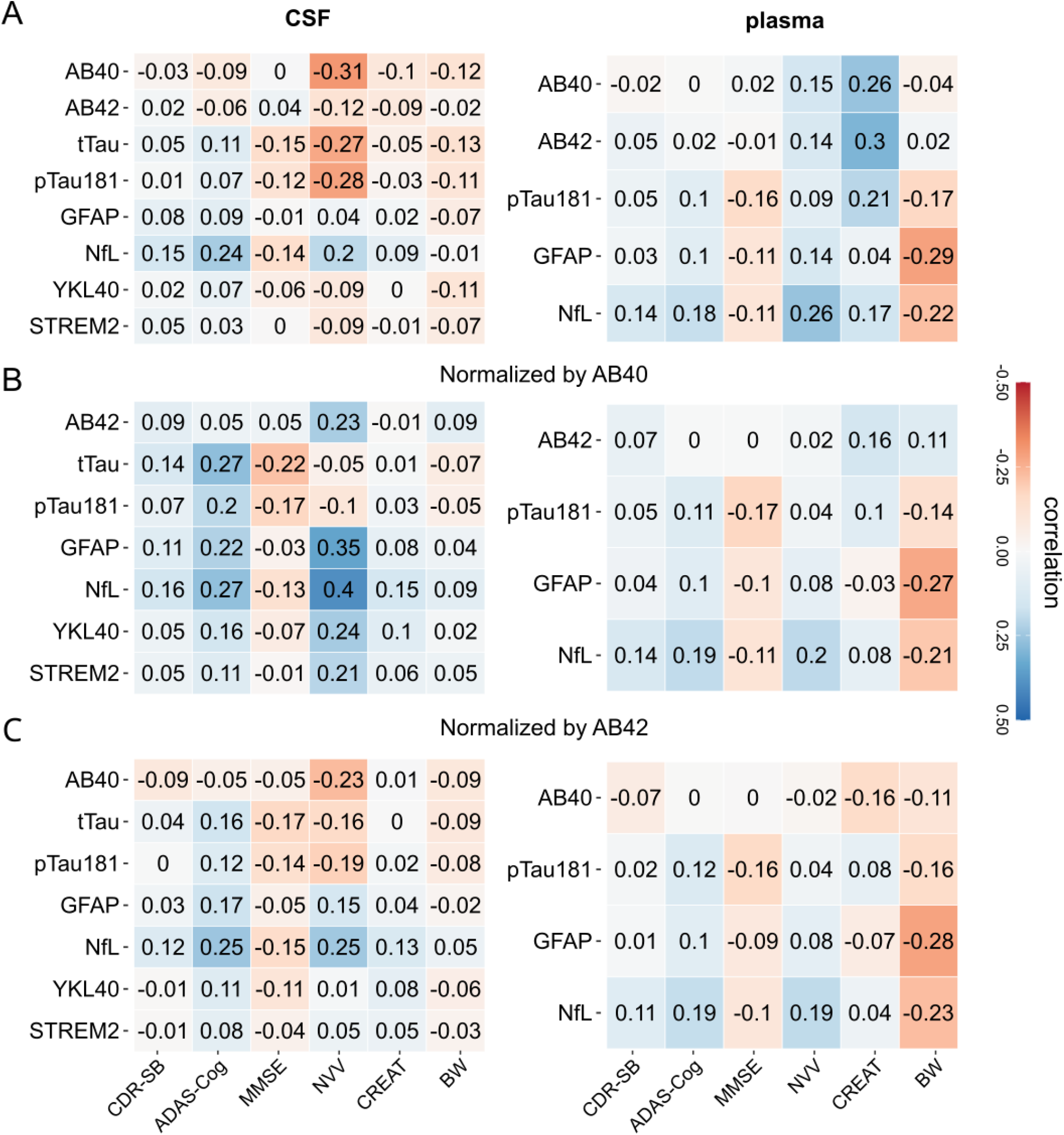
same as Figure 4 for CREAD dataset.

**Supplementary Figure 5:**
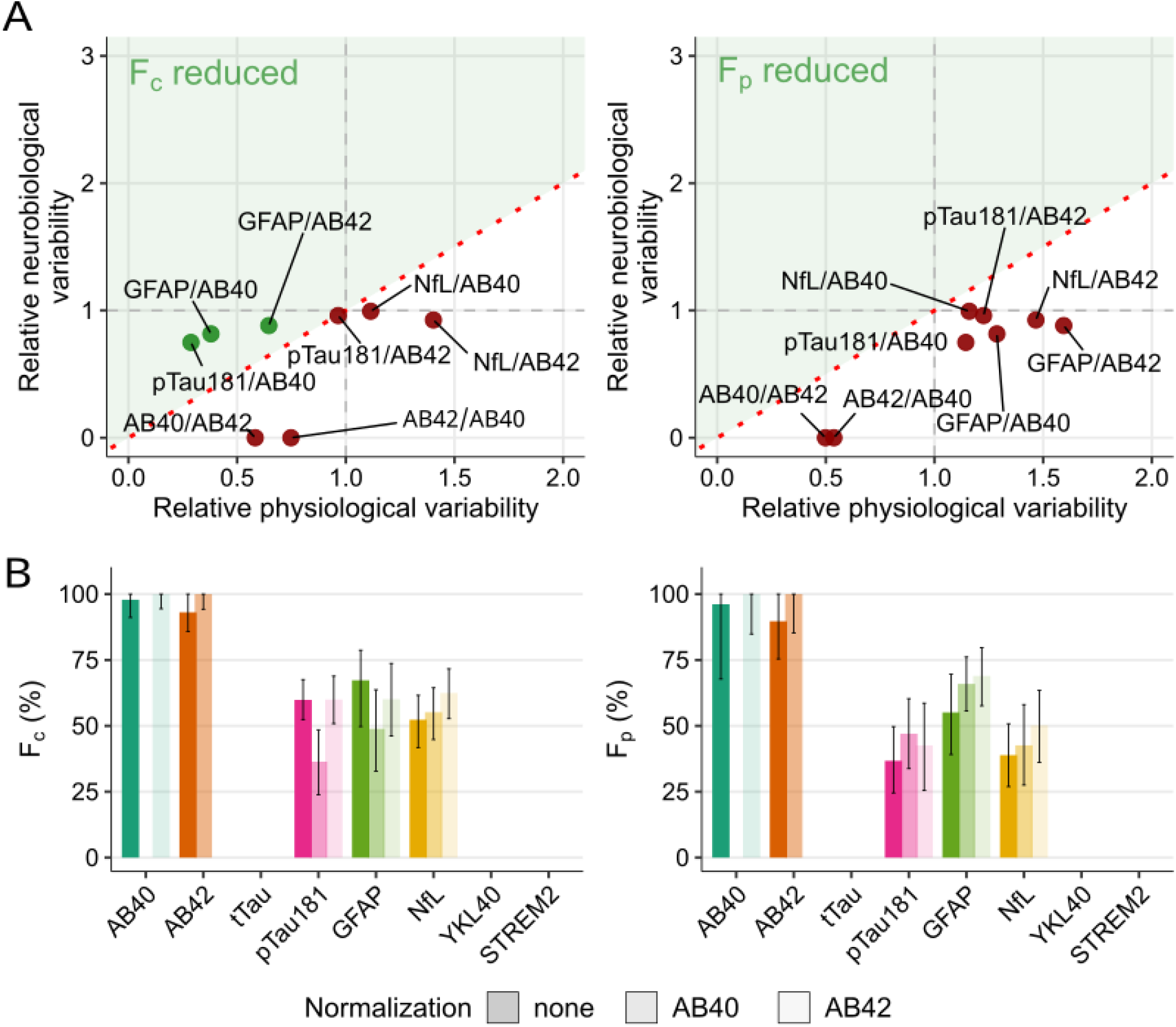
same as Figure 5 for CREAD dataset.

**Supplementary Figure 6:**
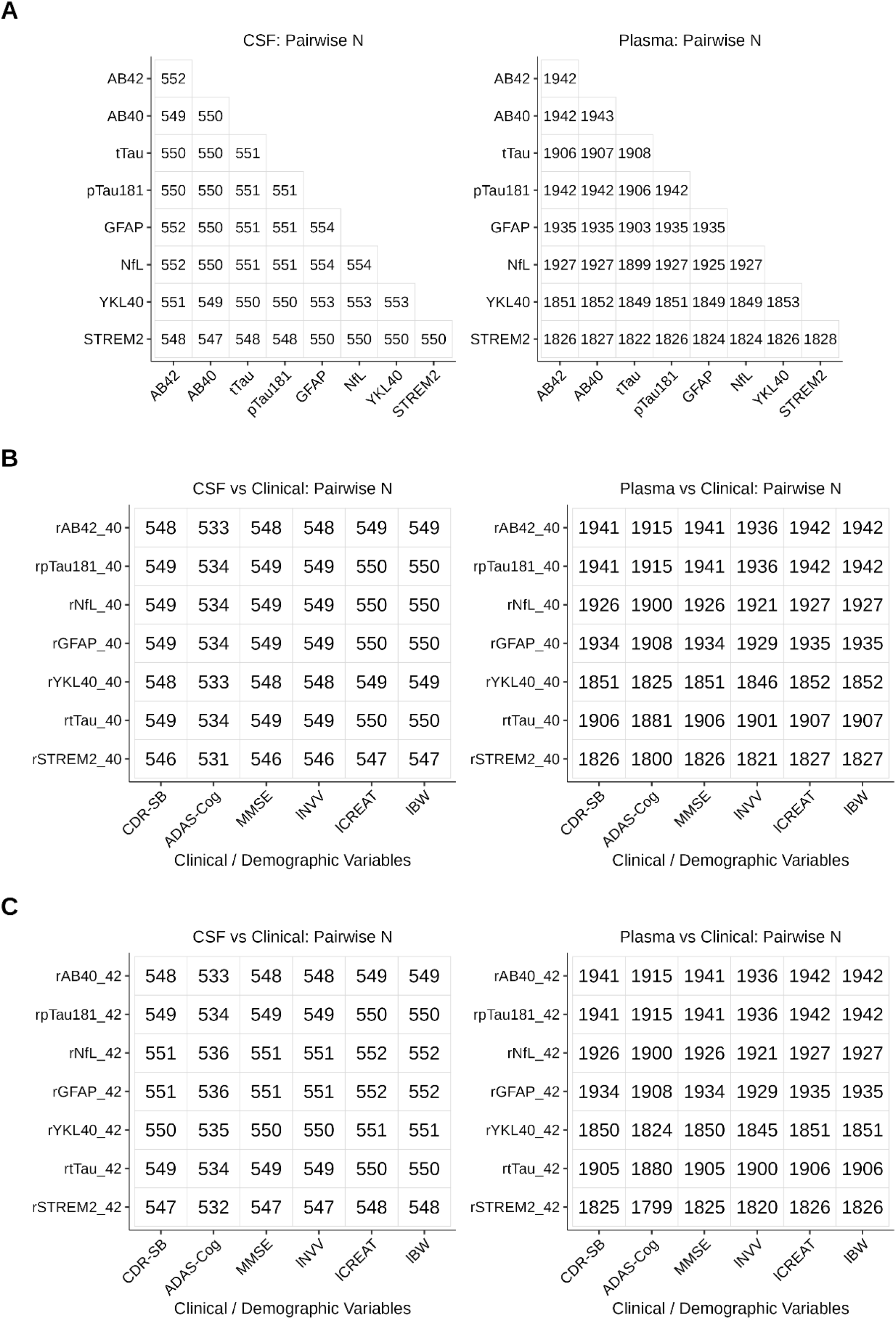
Number of individuals in the corresponding heatmaps of Figure 4 in the main text. Data: GRADUATE

**Supplementary Figure 7:**
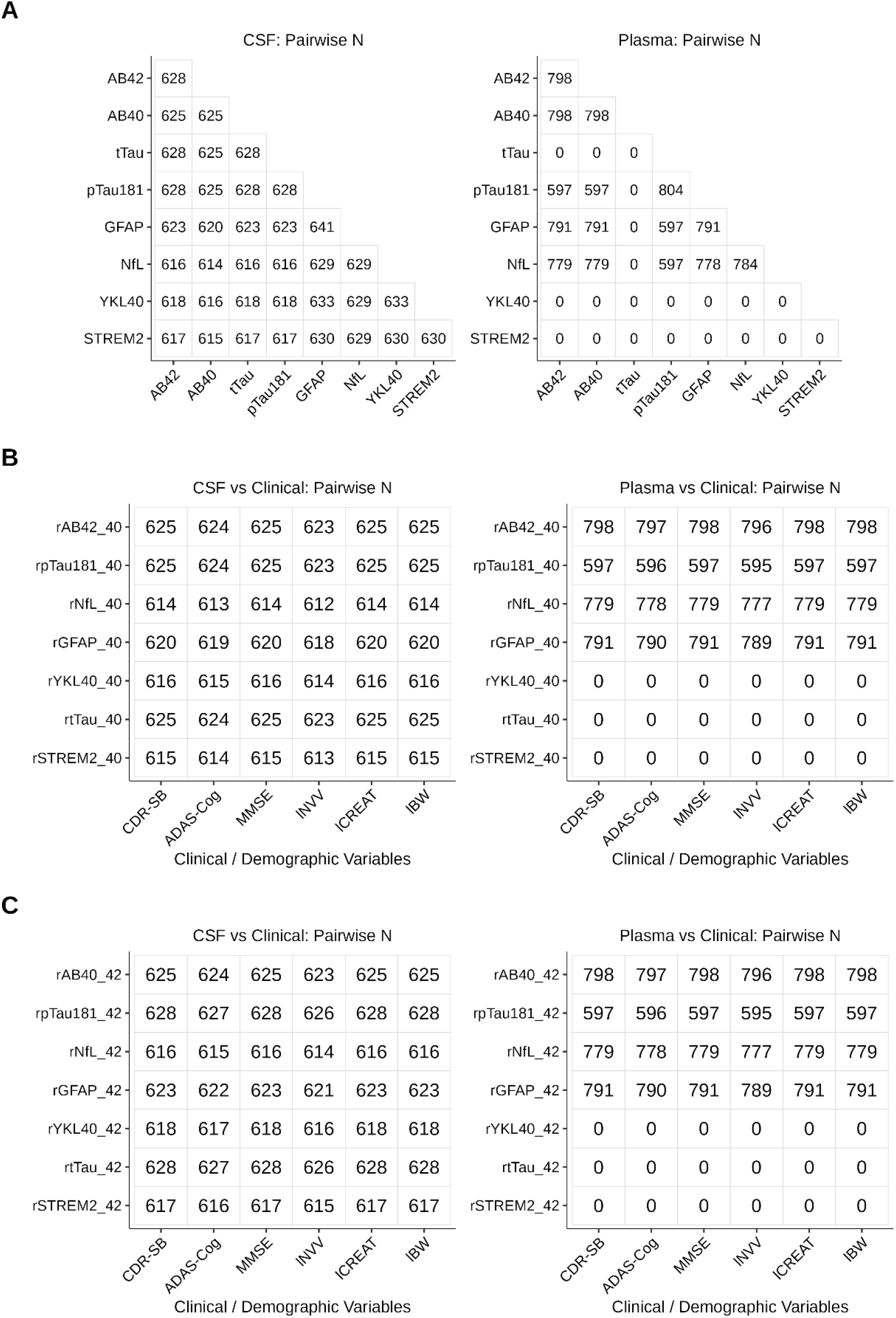
Number of individuals in the corresponding heatmaps of Supplementary Figure 4. Data: CREAD

**Supplementary Figure 8:**
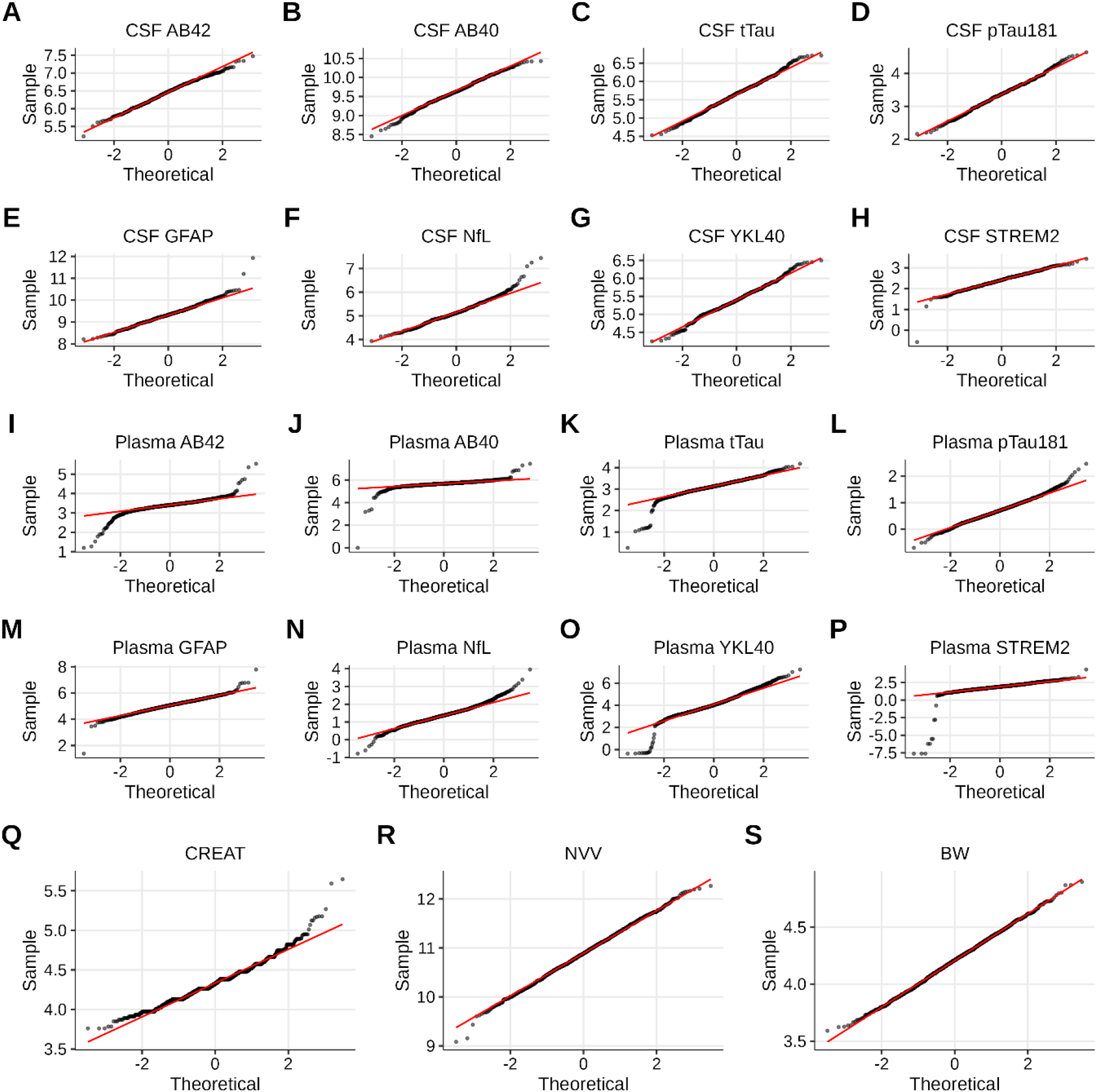
QQ plots for log10 transformed biomarkers and covariates. Data: GRADUATE studies.

**Supplementary Figure 9:**
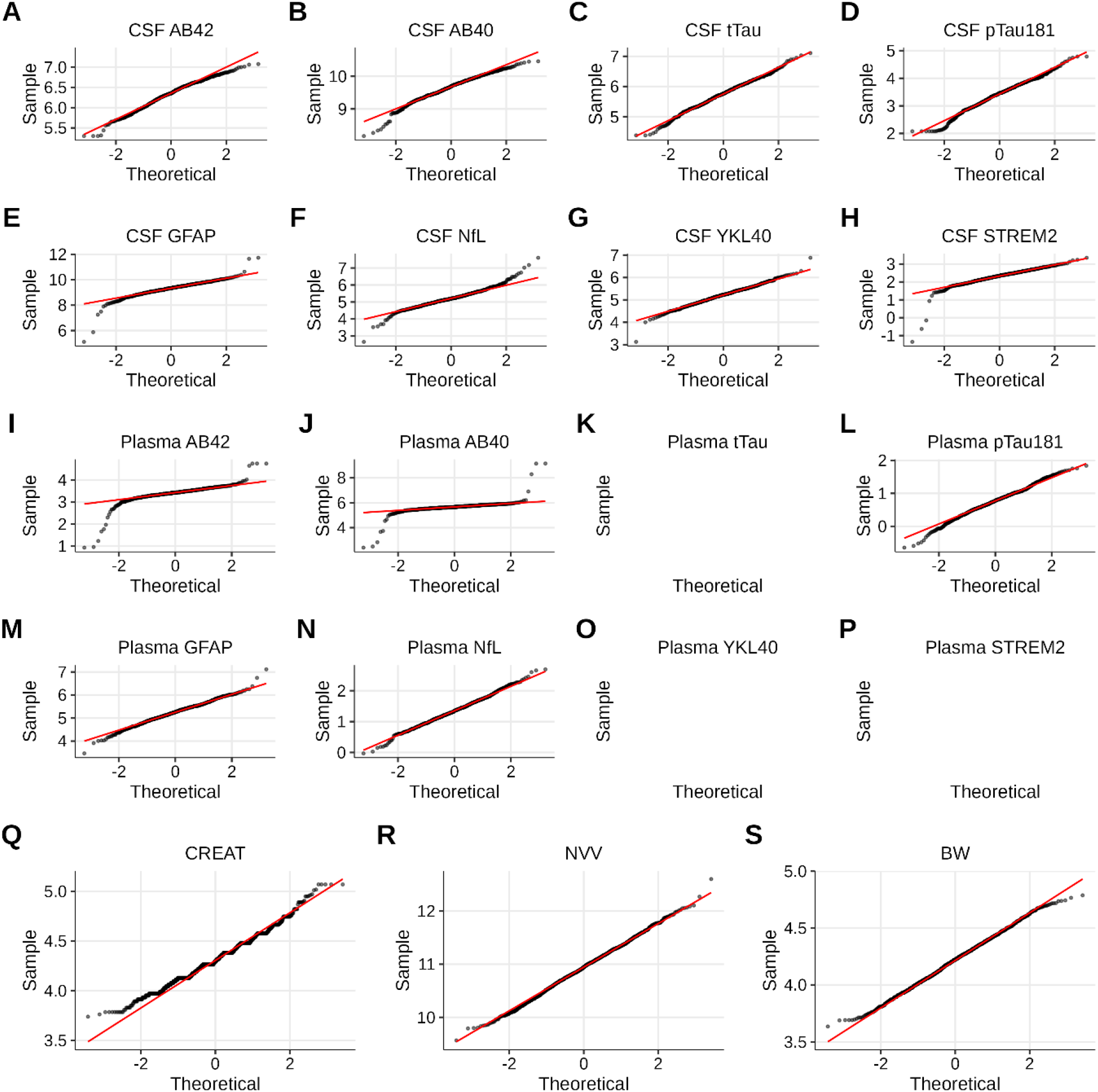
Similar to Supplementary Figure 8. Data: CREAD studies.

